# No time to waste: A synthesis of evidence on time reallocation following water, sanitation and hygiene interventions

**DOI:** 10.1101/2024.12.18.24318821

**Authors:** Hugh Sharma Waddington, Sarah K Dickin, Kishore Basak, Saranya Mohandas, Biljana Macura

## Abstract

Large amounts of time are wasted globally by households who need access to improved water for domestic uses and sanitation. The burden of inadequate access mainly affects women and girls in low- and middle-income countries. We conducted a systematic review and meta-analysis based on evidence mapping initiatives. The evidence synthesis found mean reductions of 15 minutes per trip for water supply, and 3 minutes per trip for sanitation interventions, adding up to around 8 hours per week and 3.5 hours per week respectively at the household level. Time savings from improvements in water supplies were very large, whether improved water supplies were provided at the household or community level. In contrast, studies on water treatment reported small time increases, and there were no studies that reported time following hygiene interventions. We found limited evidence on time reallocation to other activities, but disaggregated data showed girls were significantly more likely to attend school following WASH interventions. This policy-relevant evidence synthesis contributes to the case for increasing investments in appropriate water supply and sanitation interventions.

## 1. Introduction

Reducing the time needed to access water for domestic use has long been an aim of development interventions for health, social and economic reasons (1–3). Reduced water hauling time means decreased risks of water carrying injuries and assault (4–6), and this reduction in drudgery can also mean better nutritional status (7). Time savings can also come from avoided infection (e.g., costs of looking after sick children, which might affect the main carer or an older sister). In addition, economists have put an estimated value of 50 percent of after-tax wages for time saved from informal activities such as collecting water (8).

Despite the negative societal impacts, large amounts of time continue to be used for water-related work, as 1.8 billion people collect drinking water from supplies located off premises, a responsibility falling to women and girls in seven out of ten households according to the Joint Monitoring Programme (JMP) for water supply, sanitation and hygiene (WASH) (WHO/UNICEF, 2023). Included in the JMP definition of “drinking water” is water for all domestic purposes, including water for washing and other domestic hygiene practices, which comprise the majority of domestic water consumption in litres per capita per day. Compared to water, empirical studies of time taken to access shared sanitation facilities or find safe places to defaecate in the open have been overlooked until fairly recently. Even when distance to the facility is minutes from the home, time accumulates for accessing water and sanitation due to multiple trips needed per person each day, especially for women who are more likely than men to need to access defecation areas away from the home. This challenge most affects 419 million people globally who have no sanitation access at all, and who therefore practise open defecation, and over half a billion people who share sanitation facilities with other households (WHO/UNICEF, 2023). Similarly, little attention appears to have been given to time required for accessing and using hygiene facilities.

The allocation of time to WASH activities is a key manifestation of WASH-related inequalities. The hours people, usually women and girls, spend unpaid collecting water for domestic use each day (10) and using sanitation services (11) diminishes the availability for more economically and socially productive activities or leisure. The burden may involve time spent travelling long distances due to a lack of nearby water sources, travelling to secondary water sources when primary supplies dry up seasonally or due to increasing demand or unreliability of supplies. Even where water sources are nearby, long queue times are frequently a challenge, particularly in unserved urban areas, requiring substantial round-trip time from the place of water use. Time is also spent waiting for water deliveries in areas with unreliable service provision, and residents may be forced to make time decisions about purchasing water or waiting for unreliable provision (12), or spend time travelling to make service payments to utility providers (13). In the case of children, and particularly girls, collecting water for household use has been found to take time away from attending school and other educational outcomes, resulting in long-term implications for livelihood and earning opportunities (14,15). Time is also needed to travel to sanitation facilities when they are not on premises (11). Women and girls require more time to travel to safe sanitation facilities to ensure privacy and safety from gender-based violence as, unlike men, they may not be able to safely or comfortably urinate openly, so have more frequent trips each day, and they also need to use menstrual health and hygiene facilities. Aside from productive aspects, time poverty is associated with lower well-being and physical health (16,17), providing a suite of reasons to improve access to safe WASH services.

Time to access services is also known to be an important factor in determining their convenience to users, and therefore whether WASH service improvements are likely to be taken up (18).

Hence it is crucial to understand the burden on time to understand effectiveness of WASH interventions more generally.

At the same time, it is important not to assume that participants in a particular intervention benefit without empirical evaluation. Even when actual time for water collection is reduced, there may be other unpaid responsibilities to complete inside the home, and thus it is important to understand how time may be reallocated among activities and household members. In Malawi, Van Houweling, (2016) found that women living closer to a newly installed hand pump were no longer forced to wake up before sunrise to collect water, and could participate more in market or community activities. At the same, these women could also cook ‘good meals’, regularly bring their husband bath water, and work in their husbands’ fields. Improved availability of nearby water sources may also have unintended consequences, such as increasing quantities of water used which may continue or even worsen the burden of water-related work. Narain, (2014) describes the way that shorter and easier distances to collect water meant that activities done publicly at *bawdis* (i.e. stepwells), such as bathing, were instead done at home.

This meant that women were then expected to carry water home for bathing purposes. In Ghana, Arku, (2010) found that women were able to spend more time in economic activities after a water supply implementation, but men also spent less time assisting with other domestic chores such as bathing children (since less time was needed for women to collect water). In addition, there has been little attention to the ways that some common WASH interventions improve domestic hygiene, such as hand-washing promotion, or to treat water, may impose greater time burdens on those responsible, usually women and children’s carers. Little is known comparing how much time is gained due to reduced illness with time use for intensive hygiene activities. Inequalities can arise in new water schemes that reduce travel time for some but not others, especially those who are unable to afford to pay for it (22). Beyond the economic implications of time use, some researchers have reported positive social benefits of water collection or trips to practice open defecation, particularly in countries with limited opportunities for mobility outside households for women (23). A more holistic picture of how interventions impact travel time and time allocation, and how they differ in different contexts, is an important step towards better evidence-informed and gender-responsive WASH programming.

While a growing number of primary intervention studies include travel time as an outcome (24,25), we are not aware of any published systematic reviews of travel and access time or alternative time use related to interventions that aim to provide improved water supplies, water treatment and storage, sanitation and hygiene. The aim of this study is to evaluate time savings related to WASH interventions in low- and middle-income countries (L&MICs). By focusing on intervention studies from the published and unpublished literature, the synthesis offers novel policy-relevant evidence, contributing to the case for increasing investments in acceptable water, sanitation and hygiene technologies (26).

## 2. Background: methodological approaches for measuring time

There has been growing interest in analysis of time use, which requires good quality data. Several countries collect their own travel time and time use data (e.g., Ethiopia Time Use Survey 2013 (ETUS), Peru’s Encuesta Nacional de Uso del Tiempo, South Africa’s Survey of Time Use). The apparatus to monitor reported progress on water collection times at household level has been in place in most countries since the Demographic and Health Survey (DHS) included a question on the time taken to “go there, fetch water, and come back” in Phase II (1988–1993) (Institute for Resource Development/Macro International, 1990).

There are different approaches available to measure time use, which have implications for the type and quality of information generated. Specifically, data on time can be collected in several ways: direct observation of respondents, time diaries, self-reporting during a survey, or use of technologies or devices (27). These approaches have advantages and disadvantages related to costs of data collection, recall bias, ease of use and ethical issues (Table 1). Measurement of time allocation is common in the water sector (less common in work on sanitation), recall methods being most frequently used. Surveys may ask questions regarding break-down of time daily and weekly for different water and sanitation-related tasks, such as the number of trips daily or weekly, time spent travelling versus queuing, time spent on water collection for different purposes. JMP defines improved drinking water as ‘basic’ when it requires up to 30 minutes round-trip to collect it. This is roughly the individual journey time up to which basic needs for water supply can be reasonably met (3,28). Even so, a measure of the total time used to fetch water per household day might better account for multiple trips per day by large households, problems with water availability from any single source or net time required to access the new technology (e.g., filter water). In contrast, travel time for accessing sanitation or hygiene is not included in the JMP service ladder, because basic improved sanitation is defined as a toilet accessed for sole use by a single household, which would usually be in the house or yard.

**Table 1:** Different methods for collecting time data (Adapted from Masuda et al., 2014))

| *Methodology* | *Advantages* | *Disadvantages* |
| --- | --- | --- |
| Direct observation (e.g. Cairncross and Cliff, 1987; Dongzagla et al., 2020) | • No recall bias issues or requirement for same concept of time. | • Study participants may alter their behaviour in response to observation.<br>• Time to collect data and associated costs are resource intensive, limiting sample size. |
| Time diary (e.g. 24-hour time diary). | • Minimizes recall bias.<br>• Can measure primary (e.g. water hauling) and secondary activities conducted simultaneously (e.g. childcare)<br>• Can measure present company.<br>• Can measure temporal order. | • Requires literacy and numeracy to record entries, or use of pictures (Cardenas and Carpenter, 2008)<br>• Survey instruments can be difficult to administer and be overly burdensome<br>• Time may be linked to nature such as day time or seasonality (31). |
| Recall methods such as standard survey questions (e.g. Hutton et al., 2020) used household surveys to collect information on | • Simple and quick to administer. | • Can result in large recall bias, dependent on the length of recall time.<br>• It may be difficult for participants to apportion time if multiple tasks are done |
| time to access sanitation locations). |  | synchronously (e.g., child care). |
| Technology-supported methods (e.g., Global Positioning System (GPS) devices used by Crow et al., (2013) attached to jerry cans and or attached to participants used by (33). | • Can be less burdensome if data can be automatically collected and reported. | • Devices may be costly, limiting their use.<br>• There may be ethical concerns with tracking respondents.<br>• Data may be difficult to interpret, such as when someone includes other activities in the journey |

A theory of change was co-developed with stakeholders at the protocol stage that hypothesised links between gender-inclusive WASH interventions and outcomes along a causal pathway (34). To facilitate conceptual understanding of changes in time allocation following WASH improvements, Figure 1 presents a detailed causal pathway on how provision or promotion of domestic water, hygiene and sanitation services can influence travel and access time and alternative time allocation for adults, children and vulnerable groups, through improved access, reliability and quality of service, and convenience.

**Figure 1:**
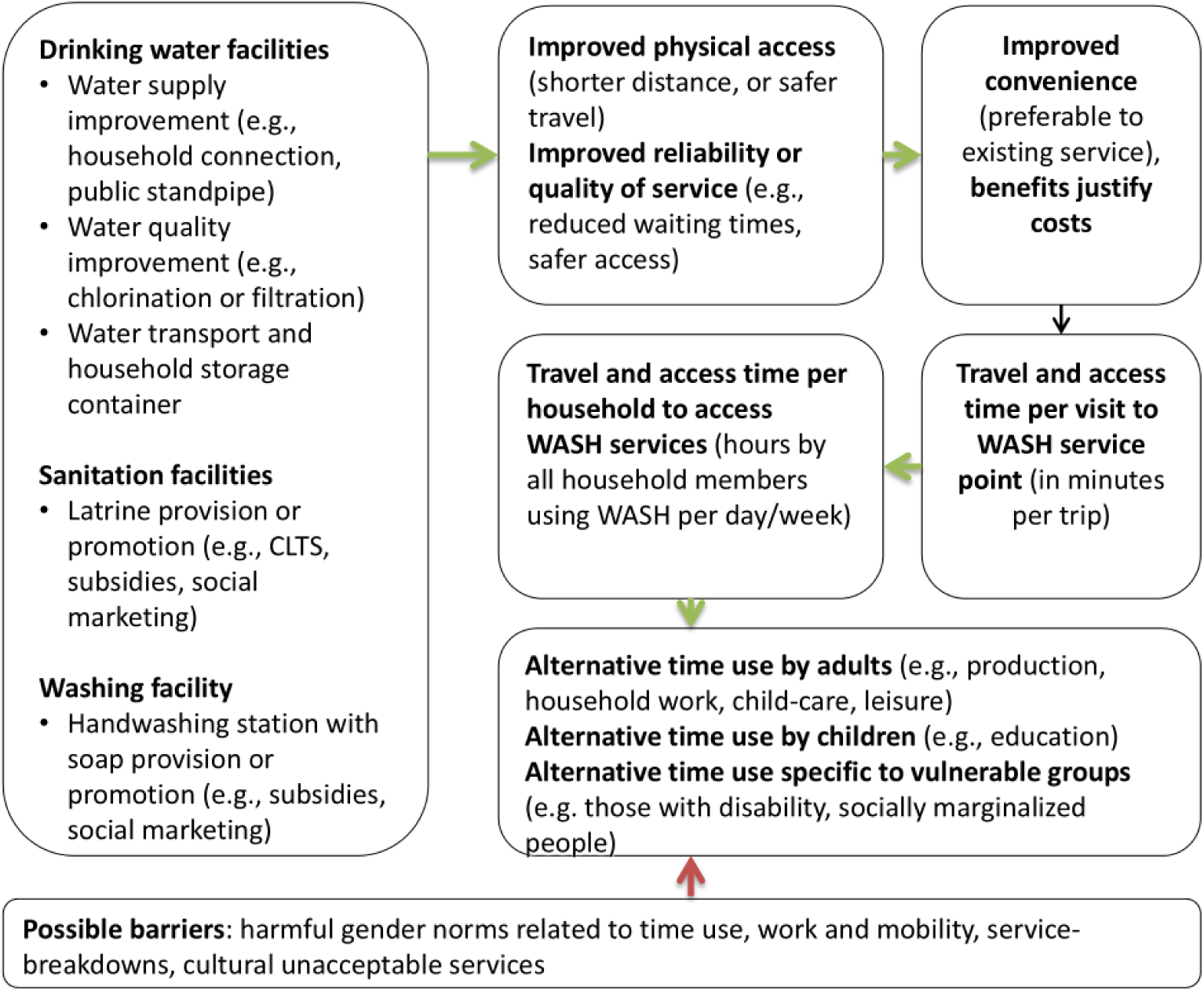
Theory of change: WASH interventions, travel time and time use

The figure shows how physical access to WASH services, reduced waiting time and perceived convenience determine time taken to access drinking water, sanitation and washing facilities, and as well as their perceived convenience, which determine use. For example, travel time savings can be made through improved tariff collection, such as through mobile (m-WASH) water tariff payments versus payment at the bank (13). Travel time may be measured per trip or per day, at the individual and household levels, so the difference in time taken per trip and the time in total number of visits per day can be substantial (e.g., David, 2004). Alternative uses of time may accrue to different groups and may be more strongly felt by those with particular needs such as people living with disabilities or the elderly, or may mainly accrue to particular household members such as male ex-water collectors (36).

## 3. Methods

This synthesis used studies from two recent mapping exercises conducted in collaboration, which systematically mapped evidence on time use outcomes of WASH interventions, focusing on general population and, in particular, women and children’s access to WASH services in households, schools and health facilities (24,25). Consequently, this review, with its particular scope, was not registered. The following sections describe the study selection process, critical appraisal and quantitative synthesis approach, drawing on the study protocol (34).

### 3.1 Search strategy and selection criteria

Searches were conducted for literature published at any time until November 2020. Electronic searches were conducted of sources of published and grey literature (including dissertations and theses), together with hand-searches of organisational websites (e.g., 3ie repositories, the World Bank, regional development banks, Oxfam, UNICEF, USAID and WaterAid). Reference snowballing was also undertaken of all included studies, relevant reviews and reference lists of books, reports and evaluations to identify studies that may not be captured in electronic searches (e.g., Briscoe et al., 1986; Cairncross et al., 1980; Chandrasekaran et al., 2022; Feachem et al., 1978; Khan et al., 1986; Saunders and Warford, 1976; White et al., 1972; WHO, 1983). Full details on search methods and sources used are published elsewhere. Stakeholders were also invited to suggest unpublished, relevant literature. Search results from bibliographic databases were managed using EPPI-Reviewer Web Version 6 (44) for de- deduplication, screening, and meta-data coding. The full search strategies and results are available in the published evidence and gap maps on which this review is based (Chirgwin et al., 2021; Macura et al., 2023).

Studies were included irrespective of publication status and electronic availability. All empirical study designs that collected data from those receiving WASH programmes, including qualitative, quantitative, and mixed-method studies, were eligible. All types of study participants residing in L&MIC contexts were eligible.

All types of interventions to promote new or improved access to WASH technologies for domestic consumption were eligible for the review, including behaviour change communication, information and health education, direct provision of WASH technologies, and economic and market-based approaches. Intervention technologies were grouped using the categories defined by Chirgwin et al. (2021). These included water supplies (e.g., piped water provision, provision of community wells and spigots, community-driven development), water treatment (chlorine provision) and water quality (e.g., information about water sources contaminated by arsenic), sanitation (e.g., promotion of pit latrines using community led total sanitation) and hygiene technologies (e.g., provision of handwashing stations) for domestic and public use (in households, schools, health facilities and community spaces). Interventions providing WASH for commercial use, such as water treatment for irrigation (e.g., Jack et al., 2019), were excluded.

Eligible outcomes were mainly time use, defined as travel time or time to access the WASH facility, and time use reallocation resulting from changes in access to WASH, such as time spent on education activities. A detailed list of definitions of each outcome is available in Annex 2. Time spent on alternative uses was defined at both intensive margin (e.g., amount of time spent at school or work) and extensive margin (e.g., school attendance and labour market participation).

Comparison conditions were existing WASH services (business-as-usual), or an intervention providing a different type of WASH technology.

Eligible studies used random assignment to WASH intervention (randomised controlled trials, RCTs) and quasi-experimental designs (QEDs), including discontinuity design, difference-in-differences applied to pre-test and post-test comparison group data, or studies using cross-sectional data with methods to address confounding such as statistical matching or adjusted regression, where there was a clear WASH intervention provided to study participants. We also included before versus after (BA, also called uncontrolled pre-test post-test only) designs. BA studies were included because time to access a WASH facility is an immediate outcome on the causal pathway, which will usually be measured with large effect (that will be larger for greater movements up the WASH ladder) and, if the period of measurement is shortly after the intervention, is unlikely to be confounded by other variables (46). We critically appraised included studies using a risk-of-bias tool (Table 2).

**Table 2:**
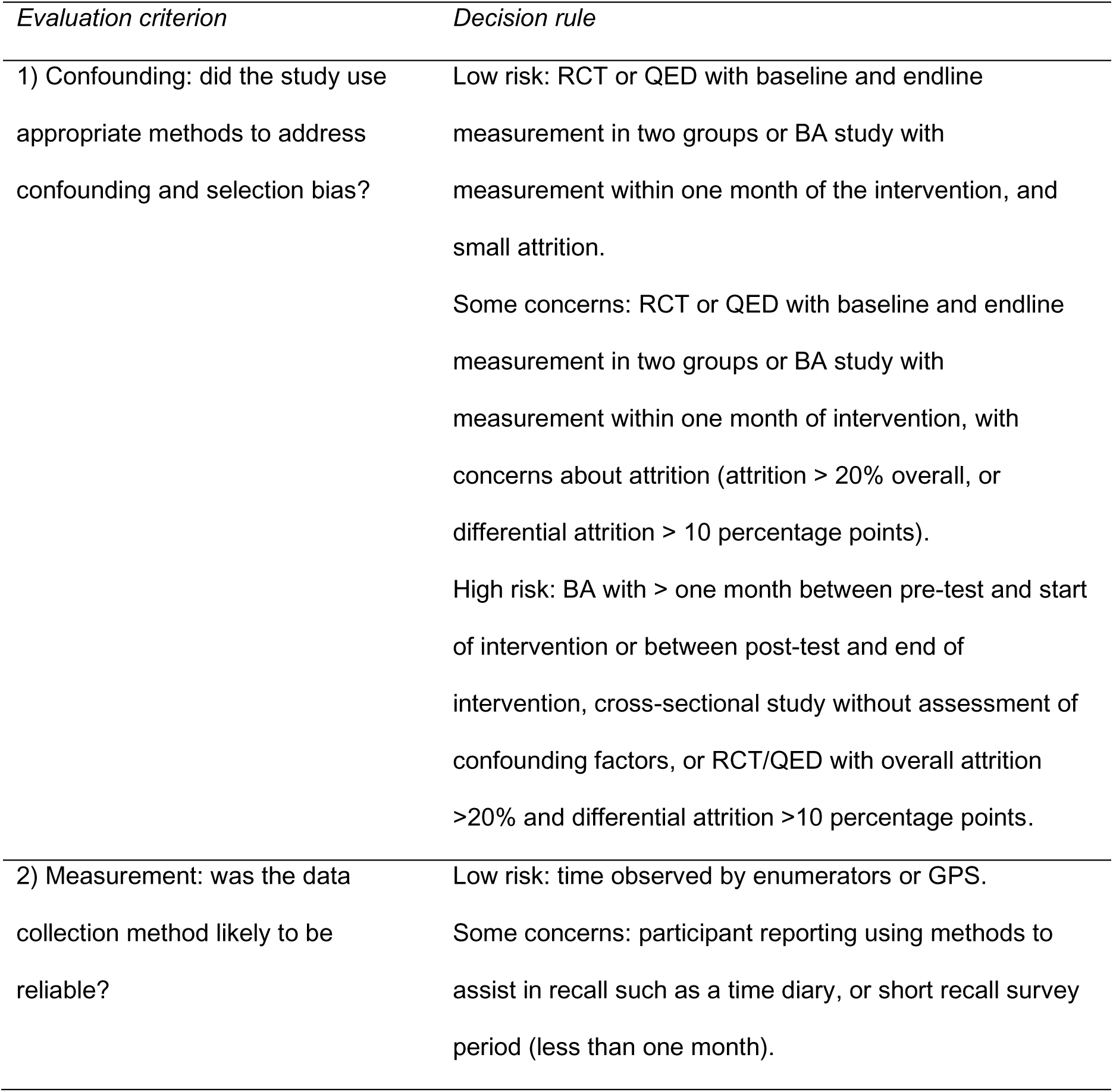
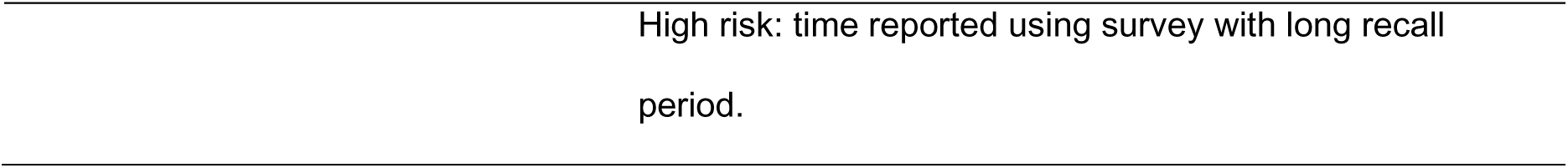
Risk-of-bias assessment criteria.

Screening was conducted in two stages by a total of eleven reviewers. First, titles and abstracts were screened together. Second, relevant records were retrieved and screened in full text. Consistency checks were performed on a subset of records at the beginning of each of the two screening stages. All disagreements were discussed in detail, with further consistency checks if the level of agreement was below 80 percent. The searches applied a combination of machine learning screening (‘priority screening’) and modelling (‘bespoke classifiers’) functions in EPPI-Reviewer Version 6 software to increase title screening efficiency.

### 3.2 Risk-of-bias assessment

We assessed the risk-of-bias using a tool we developed to evaluate the causal inferences made (internal validity) (Table 2). The risk-of-bias tool assessed two areas where threats to internal validity were considered to be especially problematic: confounding, and bias in measurement.

### 3.3 Data extraction and synthesis

We calculated time in minutes per day, hours per week, and/or using the standardised mean difference (*d*). We used a combination of narrative synthesis of time measured in natural units (minutes per trip or hours per household per week), and inverse-variance weighted meta-analysis and meta-regression analysis of time measured units of *d* (for further information on calculations made, see Annex 1). We adjusted standard errors for clustering using information reported about the numbers of clusters, average sample size per cluster and the intra-cluster correlation coefficient (ICC). Where ICC was not reported or calculable from information given in the paper, we calculated the design effect across all included studies and adjusted the sample sizes using that estimate. The design effect was usually equal to two in the studies, which meant that the variance of estimates that did not adjust for clustering appropriately was multiplied by two.

Consistency checks were performed at the mapping stage (see Chirgwin et al., 2021; Macura et al., 2023), and again at the systematic review stage on included studies. Data were collected on interventions, outcomes, effect sizes and the risk of bias by two coders independently. All disagreements were resolved through discussion.

A summary of each included individual study (including description of population, intervention, outcome) is presented in a tabular form. We generated forest plots to show the central tendency and variation in effect size estimates across study contexts. We present the meta-analysis results in natural units of time (time in minutes per trip or hours per week as reported in the studies), and used *d* to pool across outcomes in meta-regression. Sub-groups were chosen based on relevant WASH characteristics in the theory of change, such as the type of WASH technology, type of promotional intervention, type of outcome and type of population group. Publication bias analysis was also done by plotting funnel graphs and in meta-regression, following standard approaches (Higgins et al., 2023). Analyses were performed using Stata (47).

Most papers presented changes in units of time per trip or total time taken per day or week. We converted these quantities into minutes taken per trip or hours taken per week, respectively, to ensure comparability across studies. One study presented the change in time in natural logarithms (48), which we converted into units of hours per week. In a few instances, units were presented as frequencies of units of time over the sample (e.g., shares of trips taking from 0-30 minutes or 30+ minutes) (21,49–54), which we converted into odds ratios. In order to improve comparability of the estimates for the meta-analysis, we transformed all values measured in natural units and odds ratios into standardised mean differences (d-values) using formulae presented in Annex 1. Authors were contacted for additional information. For example, in one case where data on the standard deviation of the outcome were not presented in the paper, we were able to obtain primary data from the authors in order to calculate the outcome standard deviation for time spent defaecating (55).

## 4. Results

### 4.1 Review process and flow of information through review

Searches were undertaken to fully map the evidence bases on gender equality and social inclusion in WASH (Macura et al., 2023) and behavioural, health and social outcomes of WASH interventions (Chirgwin et al., 2021). Of the studies included in each review, 85 were identified as potentially relevant studies providing time-related information about access to WASH services following WASH interventions. Of these, 41 studies providing quantitative information about travel and access time and/or time use reallocation, were included in this review (Figure 2).

**Figure 2:**
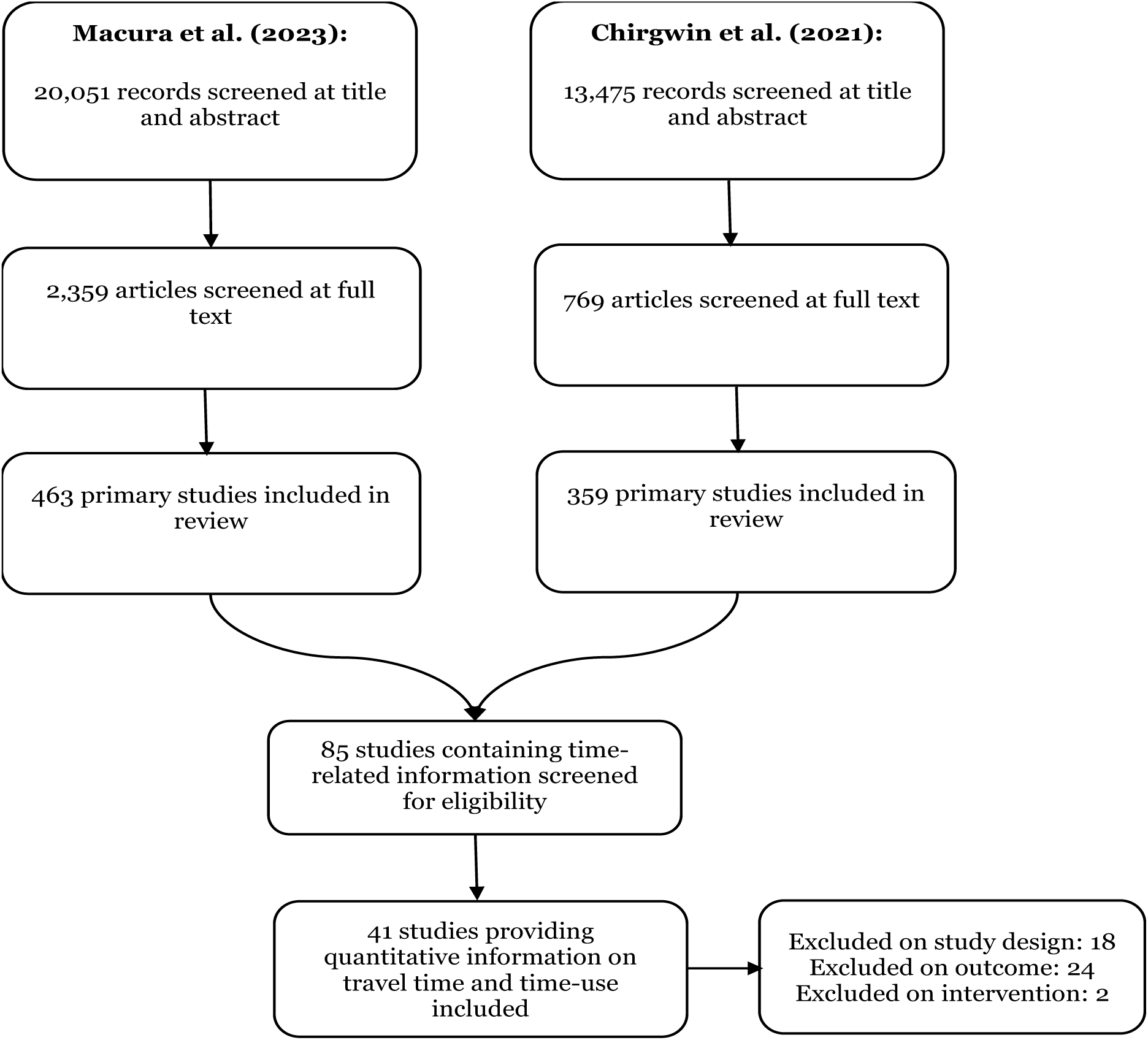
Combined study search flow diagram

Two studies were excluded because they focused on water supplies for agricultural purposes (Jack et al., 2019; Padmaja et al., 2020). Twenty-four studies were excluded on outcome, because they reported only qualitative information relating to use of time, such as the reasons for not adhering to a WASH intervention (e.g., Attala, 2019; Cronin, 2011; Hussein et al., 2017), or because they measured distance rather than time (56). Eighteen studies were excluded on study design; for example, one study reported time under the counterfactual scenario (boiling water) but not under the Lifestraw filter intervention scenario (57).

### 4.2 Description of included studies

Descriptive information about studies reporting travel or access time and time use reallocation is in Supplementary Information (Annex 2 Table A1 for water supply, water treatment and quality and Table A2 for sanitation). Below we elaborate on details of these included studies.

#### 4.2.1 Types of interventions

Various water supply interventions were evaluated (n= 33), including piped water provision to the household or yard (58), loans (59) and subsidies (Devoto et al., 2012) for piped water connections, provision of new community standpipes (e.g., Briand and Lare-Dondarini, 2017), rainwater harvesting (61), and mobile billing to improve the water supply payment process (Foster et al., 2012). Water treatment and quality interventions (n=5), including treated water sold at kiosks (Deal et al., 2020), chlorination (62,63), and provision of information about arsenic contamination in public wells (64).

Sanitation interventions (n=6) included latrine provision (WaterAid, 2015) and promotion through community led total sanitation (CLTS) (Biran et al., 2018; Cha et al., 2020; Dickinson et al., 2015; Hutton et al., 2020) and community-driven development (CDD) (Pattanayak et al., 2010). No studies measured changes resulting from hygiene interventions like the provision of washing stations or soap promotion, even though we might reasonably expect these to either reduce travel time (e.g., if washing of laundry can be done at the household) or increase it (if more time is spent washing).

#### 4.2.2 Types of populations and settings

Around half of the studies (n=18) were conducted in sub-Saharan Africa (Burkina Faso, Eswatini, Ethiopia, Ghana, Guinea, Kenya, Malawi, Mozambique, Nigeria, Zambia), 16 were conducted in South and East Asia (Afghanistan, Bangladesh, India, Pakistan, the Philippines), 5 were in Latin America and the Caribbean (Brazil, El Salvador, Guatemala, Honduras, St Lucia) and two studies was done in the Middle East and North Africa (Morocco, Yemen). The included studies of sanitation were done in rural areas in Ethiopia, India, Malawi and Zambia. The studies of water supply were also largely done in rural areas, although several were in peri-urban informal settlements in the Philippines (Aiga and Umenai, 2002) and Kenya (Bisung and Elliott, 2018), and one study was done in a refugee camp in Bangladesh (Sikder et al., 2020). A few studies were of urban piped water supply improvements including subsidies in Morocco (66) and loans in India (59), as well as m-WASH interventions, including mobile payment facilities in Kenya (13) and SMS notifications about water availability in India Kumar et al., (2018).

Most studies targeted participants in the general population (n=30). Cairncross and Cliff (1987) and Beath et al., (2013) measured observations among women only, while Toonen et al., (2014) only measured observations among children. Many studies measured observations among men and women separately (21,33,36,66,70,71) and children (33,55,66,70–74). Biran et al., (2018), Sikder et al., (2020) and WaterAid (2015) measured observations among vulnerable groups, including elderly people, the chronically sick and those with disabilities.

#### 4.2.3 Types of outcomes

The studies used a range of measures of travel time, including one-way travel time in the case of latrine use (55), and round-trip travel time in the case of water supply (e.g., (58), usually including wait times. Some studies measured travel time at the individual trip level (e.g., Dahl-Østergaard et al., 2010), while others summed up travel time for the whole day or more (e.g., Devoto et al., 2012). A few studies made evidence-informed assumptions about travel times by women and men; for example, in the case of (55) the authors assumed women would make six trips per day to urinate at the community latrine, whereas men, who could urinate around the yard, only needed to make a single trip to the latrine to defaecate per day.

#### 4.2.4 Types of study designs

The most common types of study design used were cross-section studies including pipeline designs (n=11 studies) (e.g., Cairncross and Cliff, 1987), uncontrolled studies with baseline and follow-up measurement (n=11) (e.g., Arku, 2010), controlled studies with baseline and follow-up measurement (n=9) (e.g., Almanzar et al., 2017) and cluster-RCTs (n=8) (e.g., Biran et al., (2018). In addition, one study used regression discontinuity design (76) and another used instrumental variables (64). All studies included in meta-analysis of time use reallocation used controlled designs, including one cluster-RCT (66), two controlled studies with baseline and endline measurement (70,74) and two cross-section (ADB, 2009) or pipeline designs (Cairncross and Cliff, 1987).

#### 4.2.5 Intervention effects

We were able to calculate 154 measures of effect on travel or access time or time use reallocation from the 41 studies that reported changes in time following a water supply and/or sanitation improvement. Where studies provided statistical information, the data were synthesised in meta-analysis; otherwise, they were synthesised narratively. Out of 154 measures of effect, only 81 included sampling and statistical information for which we were able to calculate an effect size (measured in minutes per trip, hours per week or *d*) together with its standard error.

### 4.3 Results of the risk-of-bias assessment

The risk-of-bias assessment is reported in Supplementary Information (Annex 2), including Table A1 for water supply or water treatment and quality and Table A2 for sanitation.

### 4.4 Summary information about travel and access time following WASH interventions

Table 3 presents summary information about **time to travel to and access WASH services** following drinking water and sanitation interventions. They indicate mean reductions in time spent of 15 minutes per trip for water supply interventions, and 3 minutes per trip for sanitation interventions (latrine promotion). Due to the need for multiple trips for water and sanitation each day, these add up to mean savings of around 8 hours per week following water supply interventions and 3.5 hours following sanitation interventions. Owing to the differences in types of water supply interventions and counterfactual scenarios, there is a large variation in findings for water supply interventions, ranging between 0.7 hours per week as a result of CDD incorporating general water supply projects (e.g., deep wells, water supply systems) in Afghanistan (68) and as much as 7 hours per week for CDD in El Salvador (Almanzar et al., 2017). The maximum number of minutes per trip saved was approximately one hour following installation of roof water catchments, reservoirs, public taps and community pipelines in Kenya (77). Sanitation studies were of different forms of latrine promotion (mainly CLTS), where mean weekly time savings varied between 0.5 hours caring for the sick and 6.2 hours including time savings from avoiding open defaecation and caring for the sick (Cha et al., 2020). In the case of water treatment and quality there were on average 3-minute increases in time per trip for chlorine provided at the water source in a refugee camp in Bangladesh (63), information about arsenic contaminated public wells in Bangladesh (64) and community water supply treatment and storage in India (78), the latter of which was measured at 0.5 hours per week.

**Table 3:** Change in travel time associated with water supply and sanitation interventions.

| *Intervention* | *Outcome* | *Mean* | *Median* | *SD* | *Minimum* | *Maximum* | *Num*<br>*studies* | *Num*<br>*estimates* |
| --- | --- | --- | --- | --- | --- | --- | --- | --- |
| Water supplies | Minutes | -15.0 | -3.2 | 21.5 | -66.4 | 0.9 | 10 | 21 |
|  | per trip |  |  |  |  |  |  |  |
|  | Hours | -7.9 | -3.9 | 10.5 | -36.1 | -0.2 | 16 | 30 |
|  | per week |  |  |  |  |  |  |  |
| Water treatment<br>and quality | Minutes | 3.3 | 3.8 | 1.3 | 1.5 | 4.3 | 3 | 4 |
|  | per trip |  |  |  |  |  |  |  |
|  | Hours | 0.5 | 0.5 | - | 0.5 | 0.5 | 1 | 1 |
|  | per week |  |  |  |  |  |  |  |
| Sanitation | Minutes | -2.9 | -3.3 | 1.5 | -4.7 | -0.2 | 3 | 6 |
|  | per trip |  |  |  |  |  |  |  |
|  | Hours | -3.5 | -4.0 | 2.0 | -6.2 | -0.5 | 2 | 11 |
|  | per week |  |  |  |  |  |  |  |

Notes: values of mean < 0 indicate reduction in time (time saving) following WASH intervention; SD standard deviation. References to each study and outcome found supplementary tables.

We also examined whether there were any differences in travel time when studies were grouped by region (Latin America, South and East Asia, sub-Saharan Africa) (Supplementary Annex Table 3). We found mean and median travel time savings per trip for water supply were larger in sub-Saharan Africa. We found on average 6 minutes were saved per trip from water supply interventions in Latin America (SD=6, range=0.5, 14; 4 estimates), 4 minutes saved per trip in Asia (SD=3, range=1, 8; 4 estimates), and 21 minutes saved in sub-Saharan Africa (SD=26, range=-1, 66; 13 estimates). When examining the median hours per week, we observed 6 hours in time saved due to water supply interventions in Latin America (SD=2, range=2, 9; 7 estimates), 3 hours in South and East Asia (SD=14, range=0, 36; 15 estimates) and 3.5 hours in sub-Saharan Africa (SD=4, range=1, 12; 7 estimates).^1^

For sanitation interventions (primarily CLTS), on average 3.5 minutes were saved per trip in South and Western Asia (SD=0.9, range=2, 5; 5 estimates) and 3 hours per week were saved in sub-Saharan Africa (SD=2, range=0.5, 6; 10 estimates). There were no estimates of time savings due to sanitation interventions measured in minutes per trip or hours per week in Latin America (Annex 2).

### 4.5 Meta-analysis of access time following water interventions

We estimated meta-analyses of WASH intervention effects on access time in units of *d* (Figure 3), and thus we were able to maximise the number of observations regardless of unit of measurement. We later present forest plots for effect sizes measured in natural units - minutes per trip and hours per week – but we do not present pooled effects in these cases since these analyses only represent a subsample of the effect estimates that we generated. The effect sizes are presented in order of publication date, in order to visually assess whether there was a general trend towards decreases in effects on travel and access time, over the course of the four decades in which studies have measured it.

**Figure 3.**
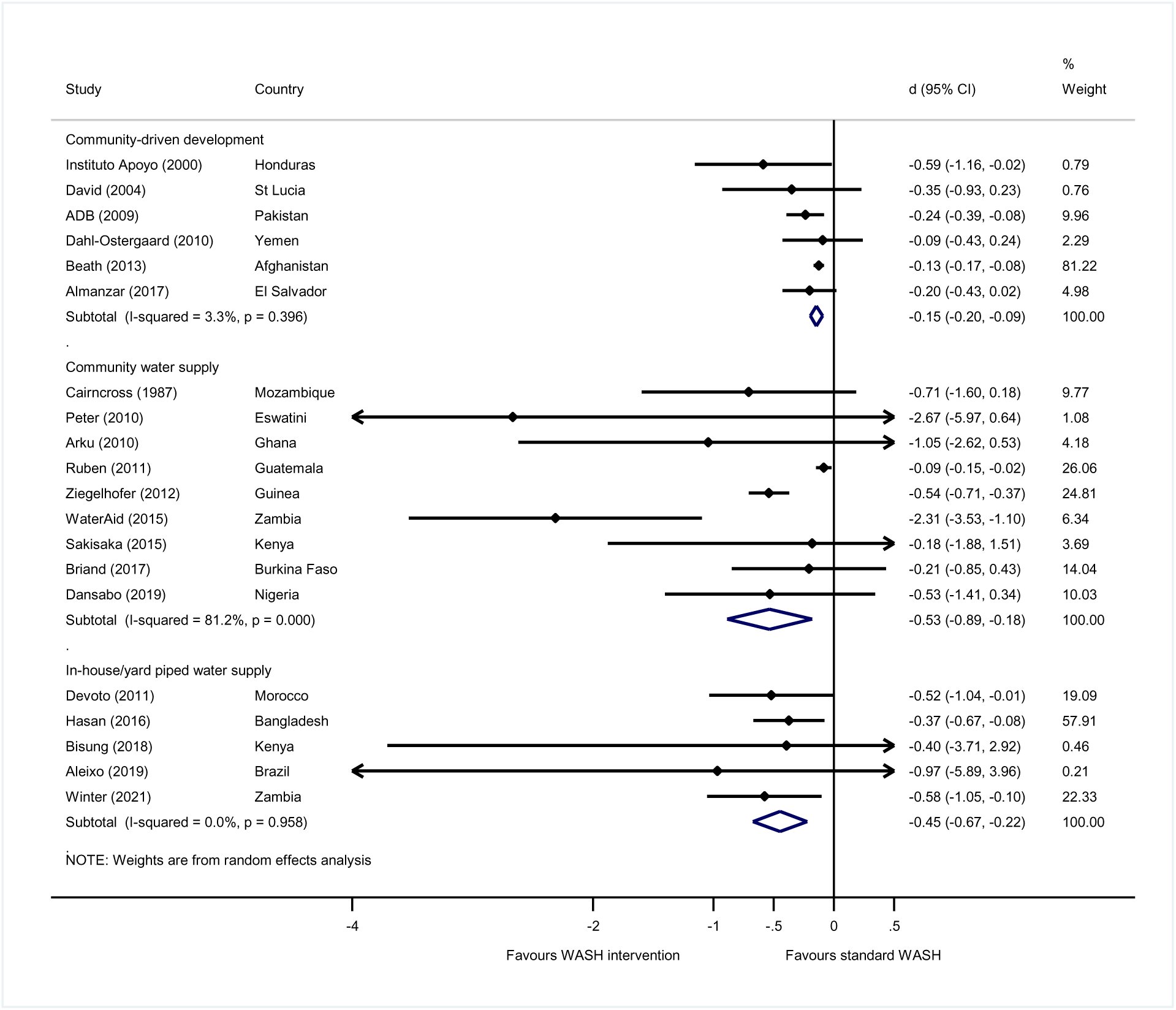
Effects of water supply interventions on travel and access time

The meta-analyses of *d* effect sizes were able to combine measures of minutes per trip and hours per week, to maximise the number of observations. An overall meta-analysis of travel and access time following water supply interventions found a moderate-sized significant pooled effect (*d*=-0.21; 95%CI=-0.29, -0.13; 25 estimates). Owing to the large number of estimates, we were able to differentiate water supply, water treatment, water quality and m-WASH interventions according to the method of promotion. We found very large effects of piped water supply provision on travel time savings (*d*=-0.45, 95%CI=-0.67, -0.22; 5 estimates) and of community water supply provision (*d*=-0.53, 95%CI=-0.89, -0.18; 9 estimates), as well as medium sized effects of community water supplies provided through CDD (*d*=-0.15, 95%CI=-0.20, -0.09; 6 estimates) (Figure 3). Statistical heterogeneity was very low for CDD and piped water, suggesting that the pooled effects are likely to be generalisable across the sample.

In the case of community water supply provision, however, heterogeneity was large relatively (I-squared=81%) and absolutely relative to the magnitude of *d* (tau-squared=0.12). While all of the interventions in this category were from sub-Saharan Africa, they measured a variety of geographical contexts, target groups and water supply starting conditions. When we examined the relative effects for community water supply interventions in circumstances where existing water supplies were unimproved according to the JMP definition – that is, were from an unimproved source like an unprotected well and/or were only accessed at round-trip journey time of greater than 30 minutes – we found larger effects (*d*=-0.82, 95%CI=-1.29, -0.36; 7 estimates) and smaller heterogeneity (I-sq=41%), than when community water supplies were provided in circumstances of existing improved water supplies (*d*=-0.08, 95%CI=-0.15, -0.02; 1 estimate from Jeuland et al., 2015). One very large effect in rural Zambia concerned access to community water supplies among vulnerable individuals, defined in that study as people with disability, the chronically sick or elderly (54). We would expect the effect of improved water supply provision for vulnerable groups with mobility needs to be greater than others.^2^

The effects of the two m-WASH interventions were heterogeneous (Figure 4), but included one underpowered but very large point estimate for an intervention that provided mobile phone water tariff payments versus payment at the bank, including wait time and return trip, in urban Kenya (13). Mobile billing may be a promising intervention to reduce travel time in urban areas, suggesting further evaluations are needed. Of the three studies of water treatment, for which the findings were very heterogeneous (and pooled effect insignificantly different from zero), one study in a Bangladesh refugee camp estimated a significant increase in access time for chlorination (63). The effects of interventions providing information about water quality also tended to increase travel time, and in only one study of information about public wells contaminated by arsenic in Bangladesh the effect was significant (64).

**Figure 4.**
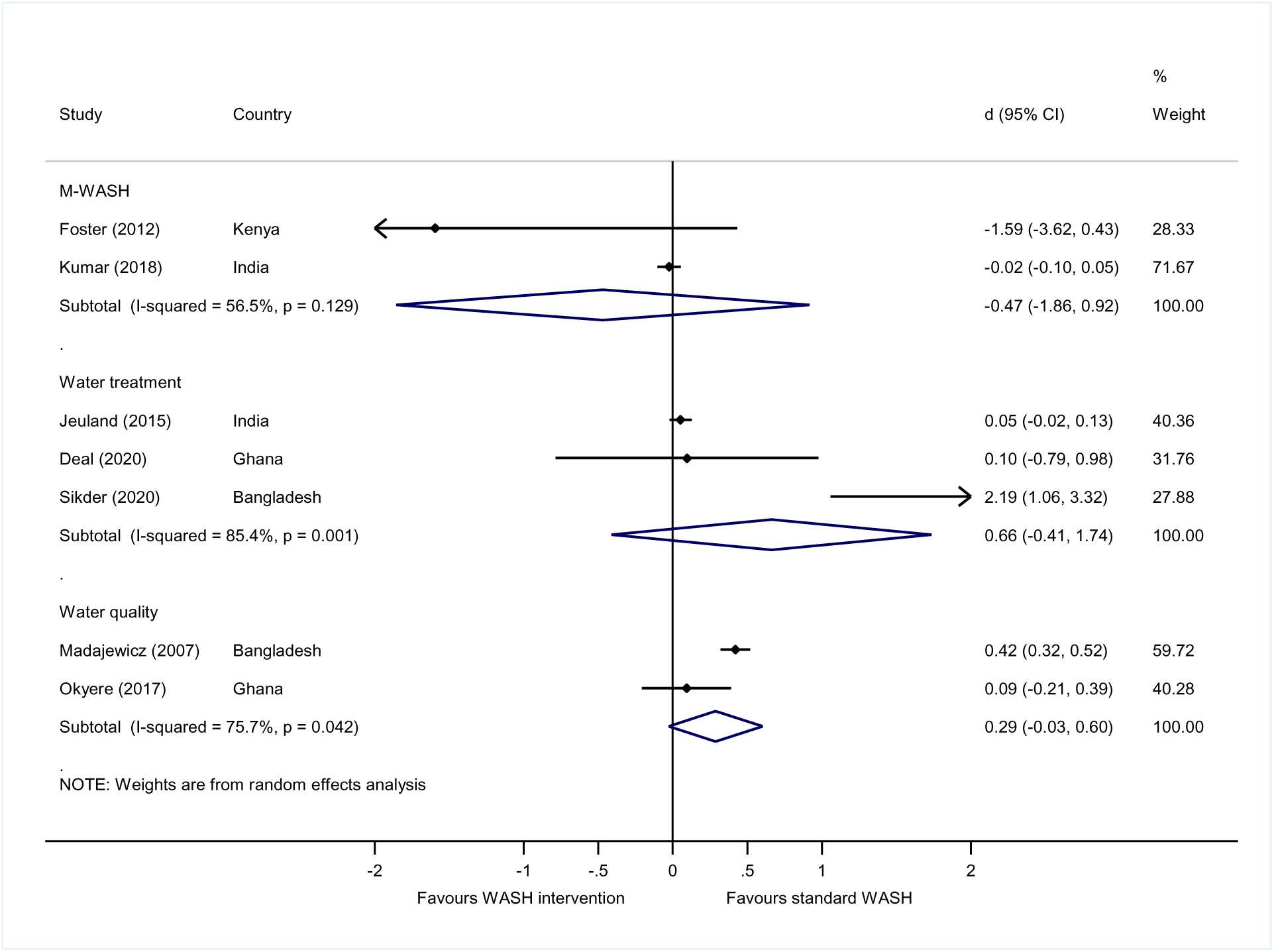
Effects of other types of water interventions on travel and access time

Turning to the presentation of findings in natural units, we found great variation in minutes per trip according to intervention type (Figure 5). The travel time saved varied considerably across individual studies, from 54 minutes saved to 4 minutes added. On average, 8 minutes (95%CI=- 13. -2; 11 estimates) were saved by water supply interventions, while 4 minutes per trip (95%CI= 3, 5; 2 estimates) were added by water treatment and quality interventions because engaging with them added to access time (e.g., chlorinating water, travelling further to an uncontaminated water source).

**Figure 5.**
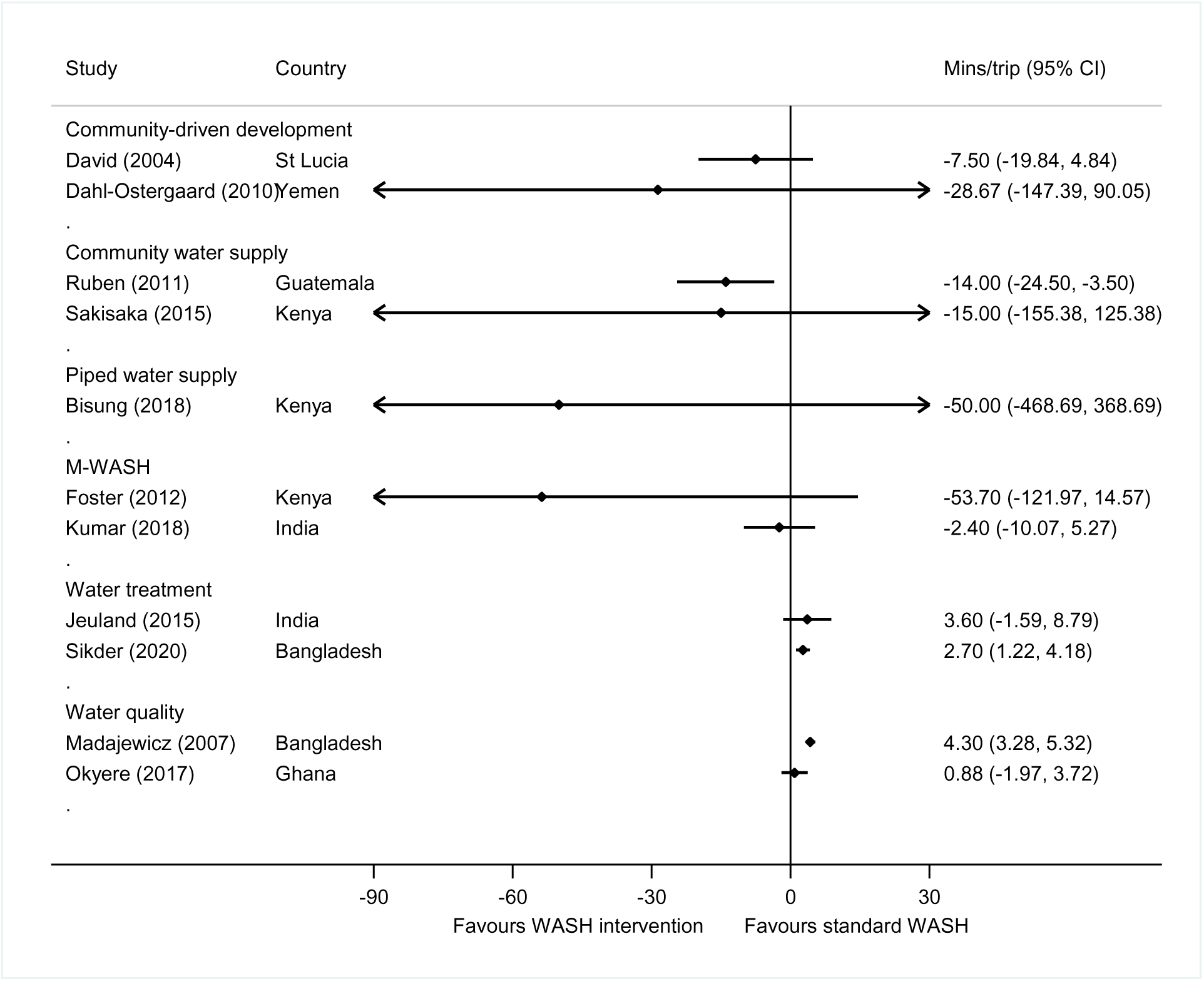
Forest plot of access time from water supply interventions in minutes per trip Note: effect sizes measured in natural units were not pooled since the analyses only represent a subsample of the effect estimates that were generated.

Regarding time measured in hours per week (Figure 6), again there was considerable variation by context, from 12 hours in Mozambique to 0.74 hours in Afghanistan. On average, 2 hours (95%CI=3, 1; 11 estimates) were saved from water supply interventions, and 0.5 hours per week were added for water treatment (95%CI=-0.2, 1.3; 1 estimate).

**Figure 6.**
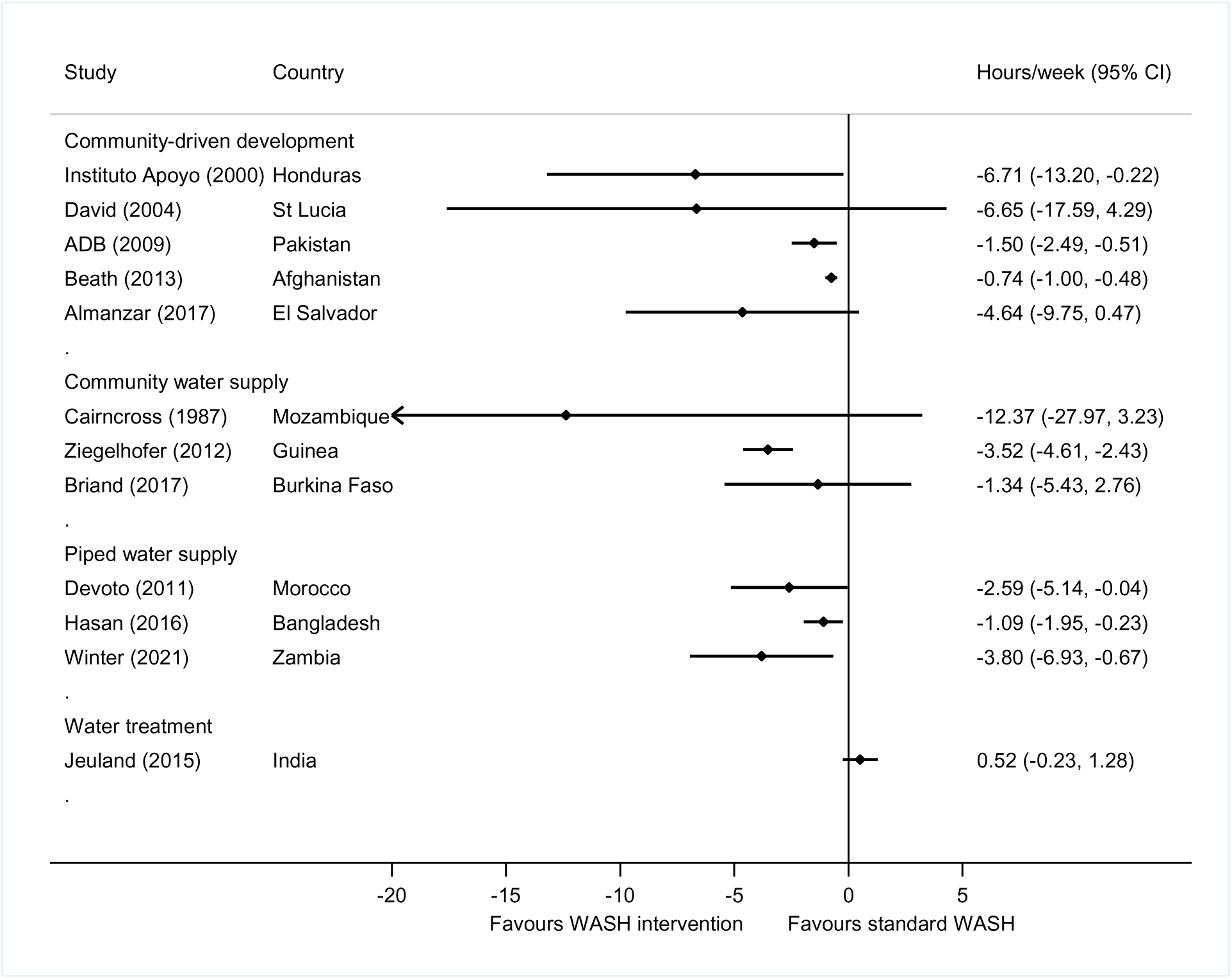
Forest plot of time-savings from water supply interventions in hours per week Note: effect sizes measured in natural units were not pooled since the analyses only represent a subsample of the effect estimates that were generated.

Three studies measured time savings for men and/or women (21,33,70) and two were conducted among women only (16,68). The meta-analysis suggested large and significant effects for women (d=-0.24, 95%CI=-0.46, -0.01; 5 estimates), but no significant intervention effects overall were found for men (Figure 7). Three further studies estimated effects on travel time of children (Table 4).

**Figure 7.**
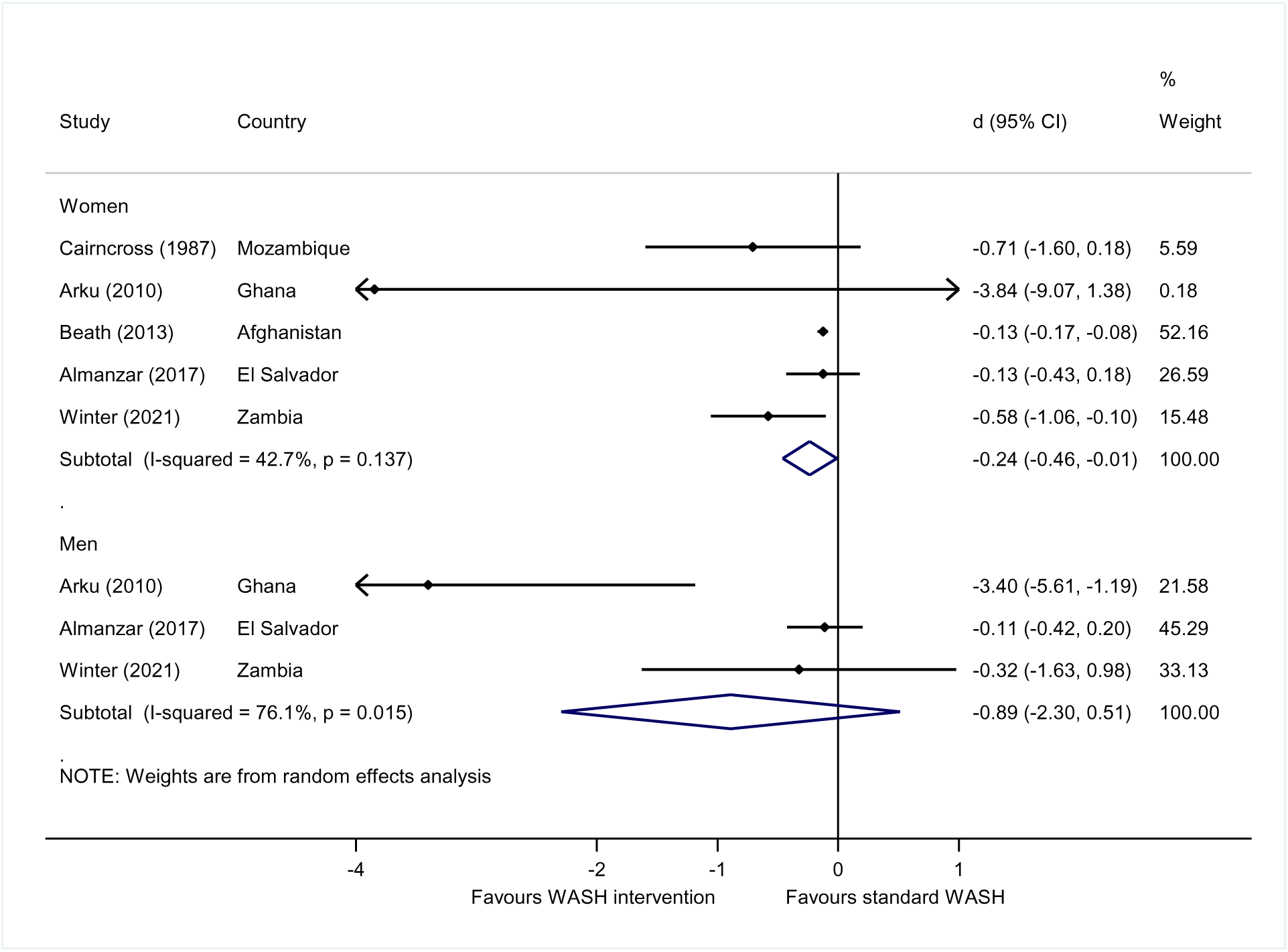
Forest plot of access time from water supply interventions by sex of adult

**Table 4.** Effects of water supply interventions on children’s travel time.

| *Group* | *Study* | *d* | *Hours/ week* | *95% CIL* | *95% CIU* | *I-sq* | *Tau-sq* |
| --- | --- | --- | --- | --- | --- | --- | --- |
| Children | Toonen et al.,<br>(2014) | -0.118 |  | -0.285 | 0.049 |  |  |
|  | Okyere et al.,<br>(2017) | -0.521 |  | -0.821 | -0.221 |  |  |
|  | Pooled effect size | -0.300 |  | -0.693 | 0.094 | 81% | 0.07 |
| Girls | Winter et al.<br>(2021) |  | -1.40 | -3.32 | 0.52 |  |  |
| Boys | Winter et al.<br>(2021) |  | -0.30 | -6.00 | 0.00 |  |  |

### 4.6 Meta-analysis of travel time following sanitation interventions

The meta-analysis of *d* for travel and access time saved from sanitation interventions, found medium-sized effects for households on average (d=-0.20, 95%CI=-0.41, 0.00, 5 estimates) (Figure 8). There was great consistency in the estimates but some estimated heterogeneity in the findings (I-squared=61%; Tau-squared=0.03). In one further study in Zambia (54), the effects for vulnerable groups were large and statistically significant (d=-1.04, 95%CI=-1.64, -0.43).

**Figure 8.**
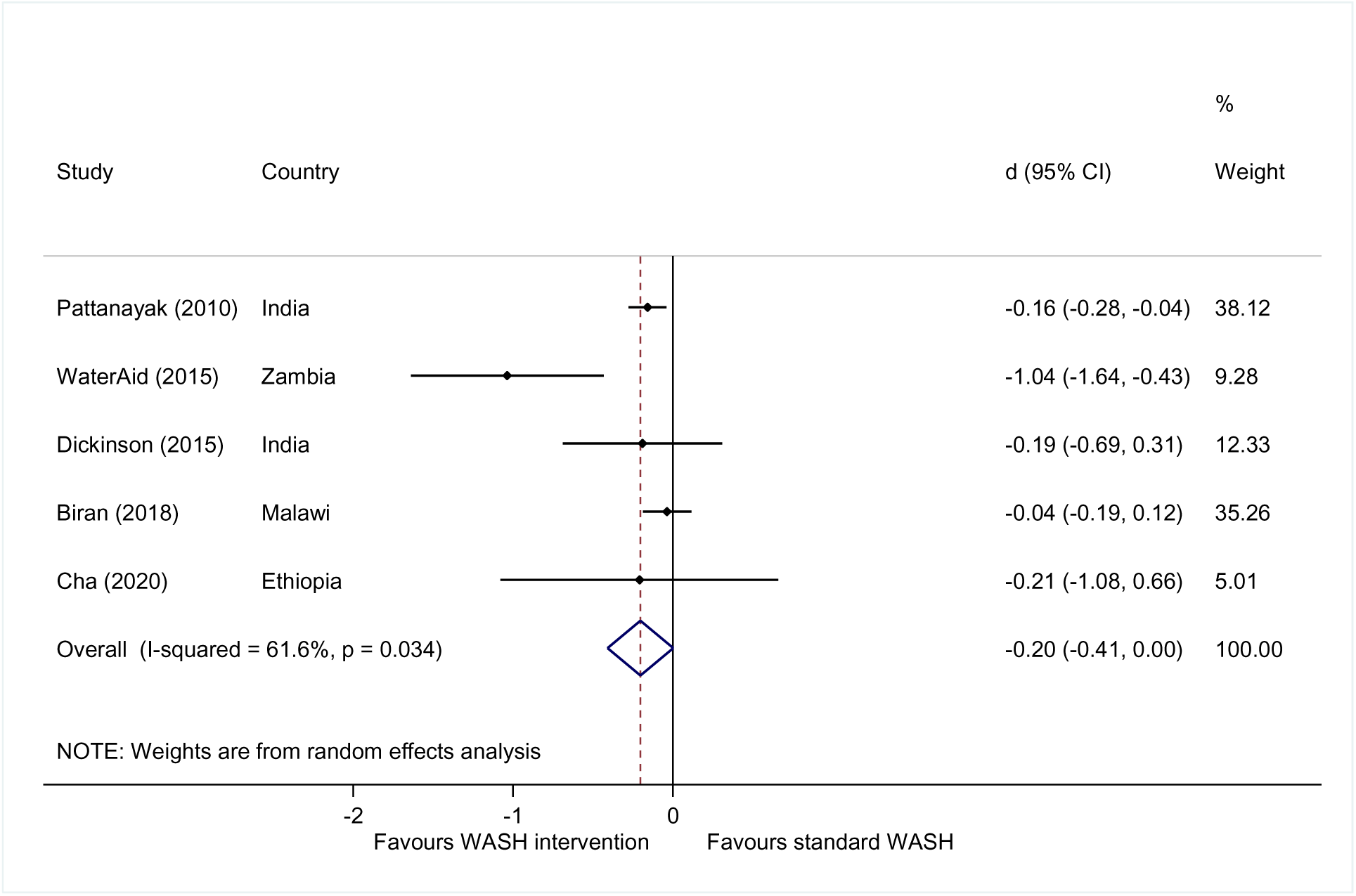
Forest plot of time-savings from sanitation interventions

Three studies measured travel time in minutes per trip (Figure 9). Two of the point estimates were from studies undertaken in India at around the same time and presented approximately 4 minutes time saving per visit to the sanitation facility. Another study of disability-inclusive CLTS in Malawi found virtually no reduction, suggesting that promotional activities were insufficient to improve access for disabled people there (75). A further study, in Ethiopia, estimated time savings of 5 hours per week per household on average (95%CI=-24 mins, 15 mins).

**Figure 9.**
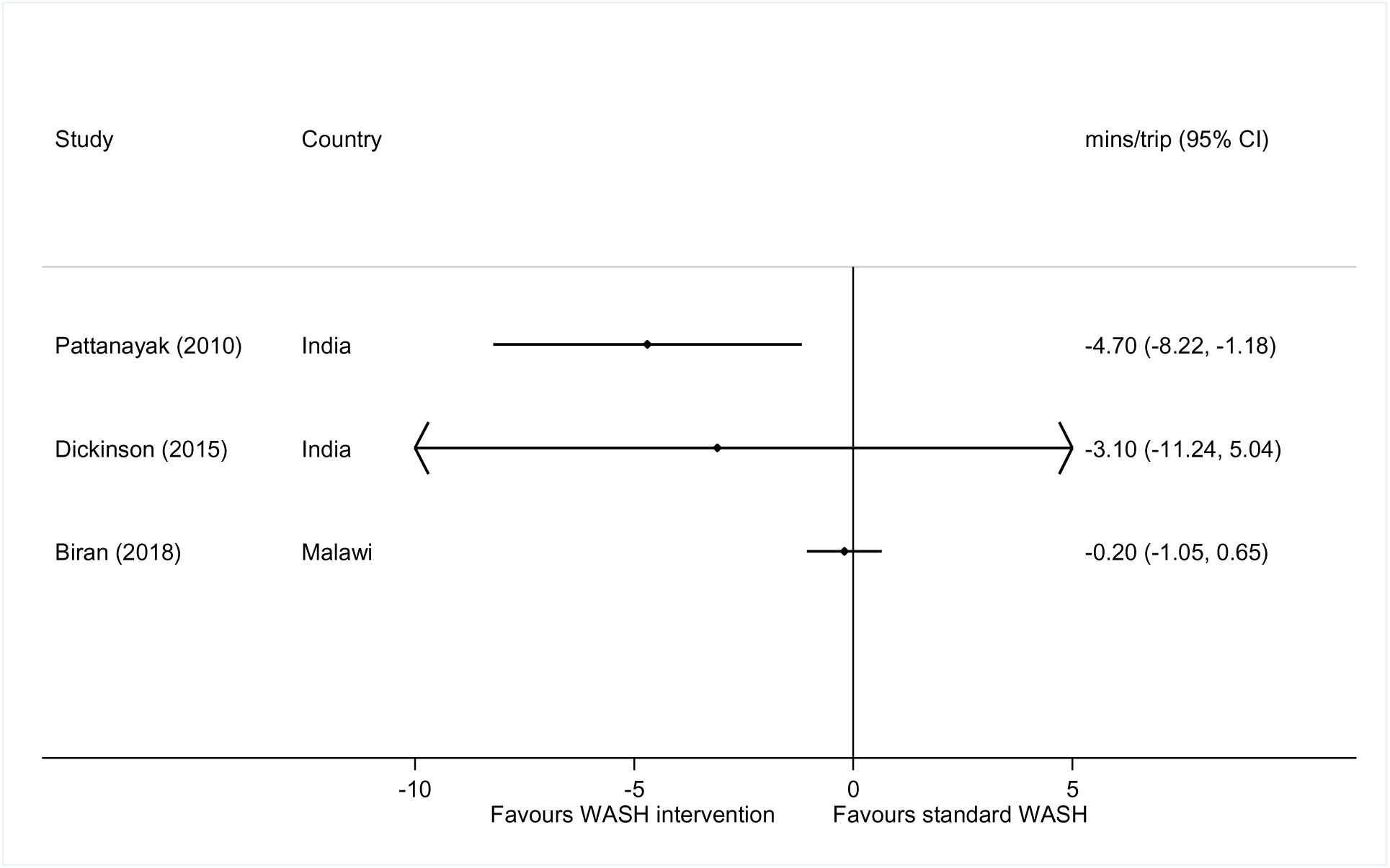
Forest plot of travel time from sanitation interventions in minutes per trip Note: effect sizes measured in natural units were not pooled since the analyses only represent a subsample of the effect estimates that were generated.

The analysis suggested women and men benefited from sanitation interventions, although only one study of sanitation disaggregated by sex (71) which reported d=-0.20 (95%CI=-0.70, 0.29) for women and d=-0.18 (95%CI=-0.67, 0.32) for men. A further two studies also reported time savings for children (d=-0.27, 95%CI=-0.66, 0.11) (Table 5), but indicated effects that were not statistically significant.

**Table 5.** Effects of sanitation interventions on travel time for women, men and children.

| *Group* | *Study* | *d* | *95%CIL* | *95%CIU* | *I-sq* | *Tau-sq* |
| --- | --- | --- | --- | --- | --- | --- |
| Women | Dickinson et al. (2015) | -0.200 | -0.695 | 0.294 | - | - |
| Men | Dickinson et al. (2015) | -0.175 | -0.670 | 0.320 | - | - |
| Children | Cha et al., (2020) | -0.383 | -0.982 | 0.217 | - | - |
|  | Dickinson et al. (2015) | -0.195 | -0.704 | 0.314 | - | - |
|  | Pooled effect size | -0.273 | -0.661 | 0.114 | 0% | 0.00 |

### 4.7 Meta-analysis of time reallocated following water and sanitation interventions

The opportunity costs of time spent fetching water and travelling to defaecate were measured as the time reallocated to other activities following WASH interventions (Figure 10). Meta-analyses were estimated for men and women separately, showing no consistent differences in time use on other activities, such as work or leisure. The meta-analysis of findings from three studies (16,66,70) reporting time use by women, found no difference overall in time use on child-care, working or leisure time by women, on average. One study measured non-significant reductions in women’s time spent doing laundry and socialising. One study – the only one that used structured observation of time use rather than reporting – suggested time available for child-care and leisure may have increased (Cairncross and Cliff, 1987). There were no differences in time spent by men following the WASH interventions (Table 6).

**Figure 10.**
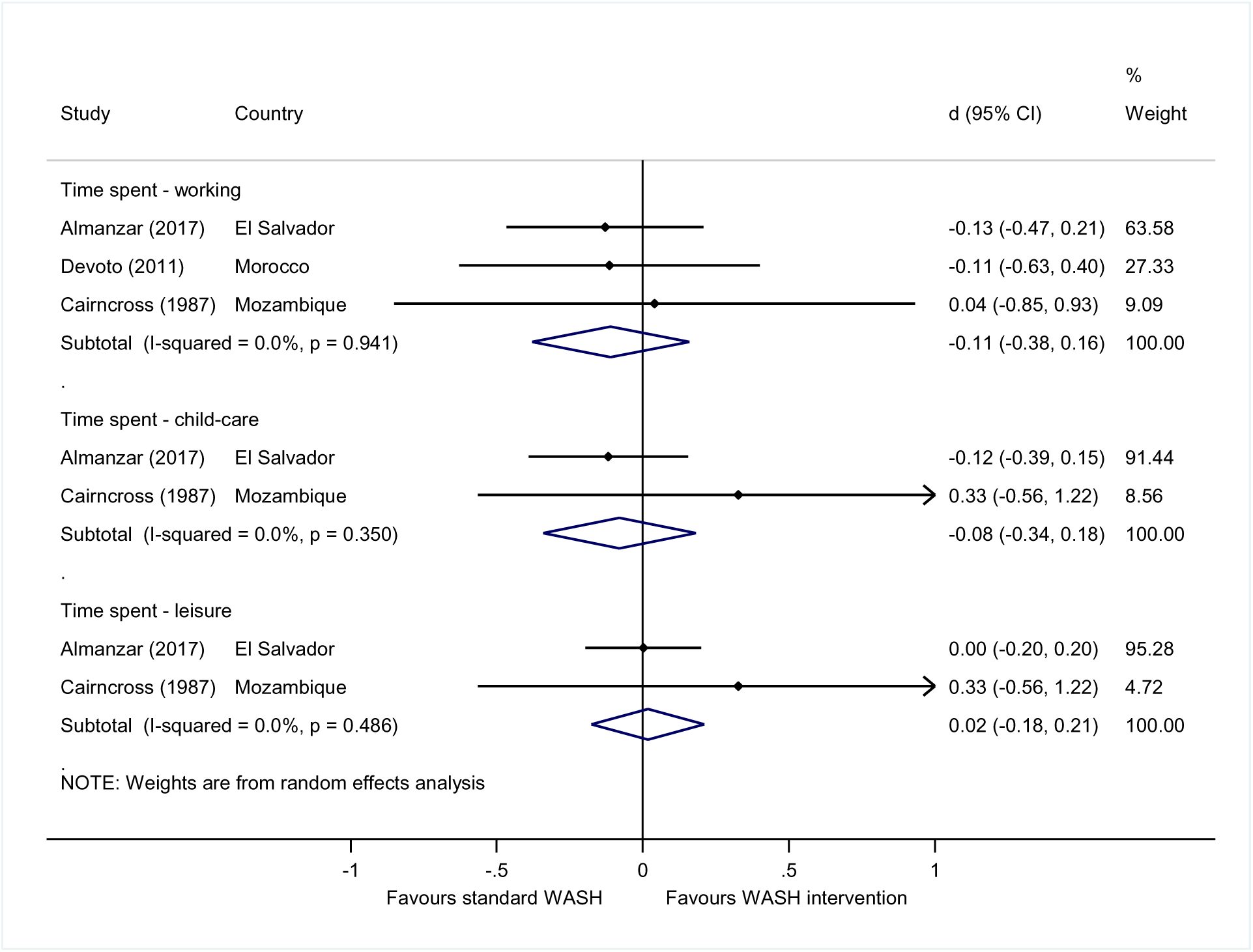
Forest plot of women’s reallocation of time following water supply and sanitation interventions

**Table 6.** Men’s reallocation of time following water supply and sanitation interventions.

| *Outcome* | *Study* | *Effect size* | *95%CI L* | *95%CI U* | *I-sq* | *Tau-sq* |
| --- | --- | --- | --- | --- | --- | --- |
| Time spent - working | Almanzar (2017) | -0.054 | -0.391 | 0.284 |  |  |
|  | Devoto et al., (2012) | 0.002 | -0.512 | 0.516 |  |  |
|  | Pooled effect size | -0.037 | -0.319 | 0.245 | 0% | 0.00 |
| Time spent - laundry | Almanzar (2017) | -0.054 | -0.497 | 0.39 | - | - |
| Time spent - social | Almanzar (2017) | 0.032 | -0.176 | 0.24 | - | - |
| Time spent - leisure | Almanzar (2017) | -0.064 | -0.267 | 0.14 | - | - |

Regarding children’s reallocation of time, the studies found medium-large sized effects of water supply interventions for girls (d=0.20, 95%CI=0.06, 0.33, 4 estimates) with no estimated heterogeneity in estimates across studies (I-squared=0%, tau-squared=0.00) (Figure 11).

**Figure 11.**
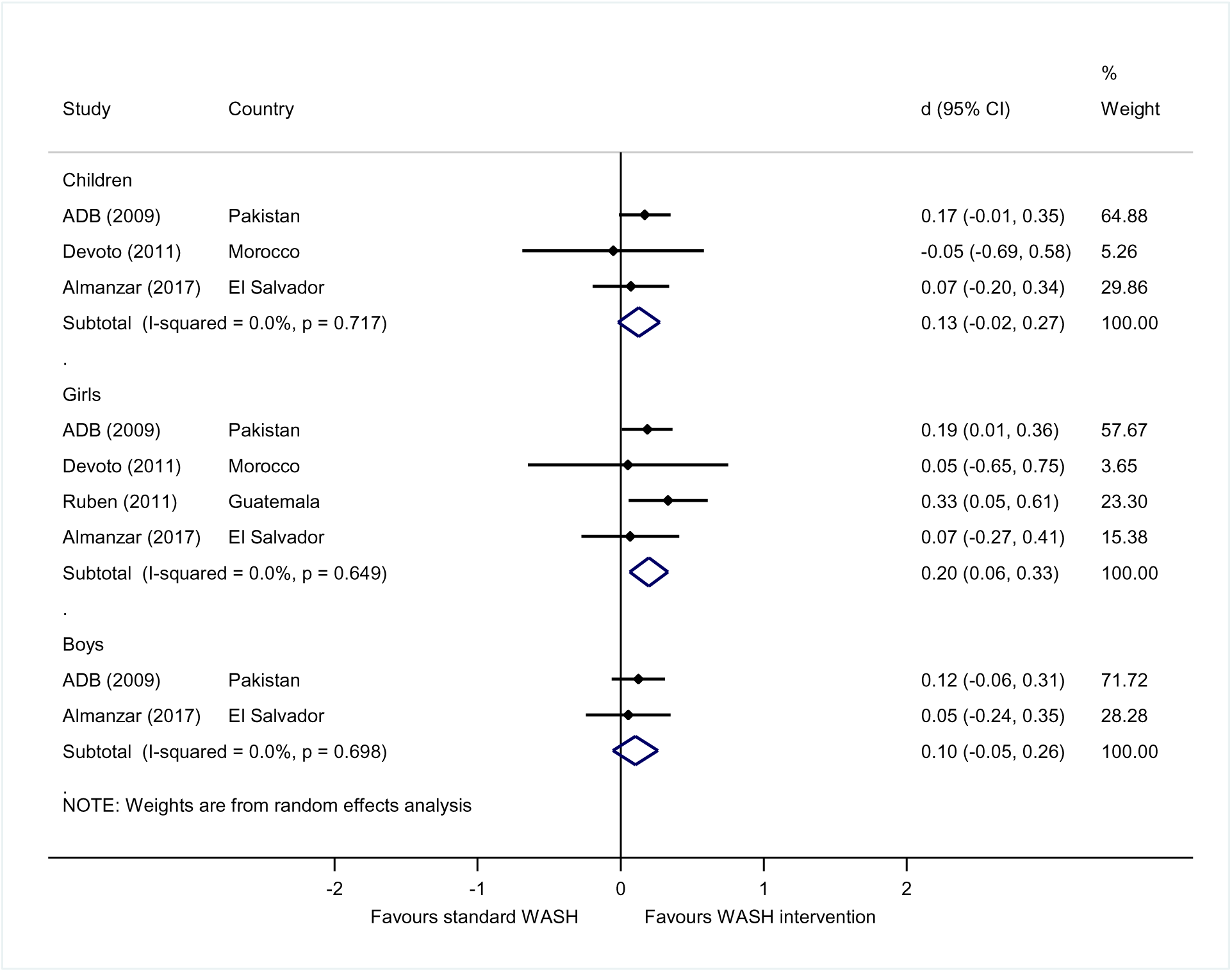
Forest plot of children’s reallocation of time for school following water supply and sanitation

### 4.8 Meta-regression and publication bias analysis

We attempted to explain the heterogeneity in findings using meta-regression, finding significantly larger effects for vulnerable groups, and smaller effects for studies assessed as at high risk of bias (Annex 3 Table A6). Publication bias tests suggested some evidence for small study effects for travel and access time outcomes (Annex 3 Figures 1A and 2A).

## 5. Discussion

We identified mean reductions in travel and access time related to water supply and sanitation interventions. With multiple trips needed per day, these add up to mean savings of around 8 hours per week following water supply interventions and 3.5 hours following sanitation interventions. Piped water supply provision had the greatest effect on reducing time for access compared to other water supply interventions, although it is important to note that, due to differences in types of water supply interventions and counterfactual scenarios, there is a large variation in findings. In the context of growing water stress, our findings have important policy implications as climate change could increase water collection time by up to 30% globally and up to 100% regionally (79)

In the case of water treatment and quality interventions, 3 minutes per trip were added on average due to time needed for activities such as chlorinating water, or travelling further to an uncontaminated water source. Although WASH practitioners promote reduced time spent on water collection, information on any additional time spent, by whom and implications on uptake including inconvenience is rarely measured. In the case of mobile billing (m-WASH), the evidence, albeit from only two studies with concerns about bias and heterogeneous effects, suggested it may be very a promising intervention to reduce travel time in urban and peri-urban areas, but further evaluations are needed.

We also found a clear trend over time in the effect sizes, as the earliest intervention studies found larger effects. Since general access to water and sanitation has improved over the decades, we expected this to be the case, assuming most of the differences in effects on time are due to pre-existing WASH service access and the degree of movement up the water and sanitation ladders. For instance, Cairncross and Cliff (1987) estimated a reduced trip time from 5 hours to 10 minutes to a water collection point in Mozambique, while Winter et al. (2021) estimated a reduction in median round-trip fetching time from 13 minutes to 2 minutes.

The results varied by region and participant group. We found that time savings tended to be larger in sub-Saharan Africa, with on average 21 minutes saved per trip compared to 4 minutes in Asia, following water supply interventions. Access to water supply is more limited in sub-Saharan Africa, where nearly half of people rely on water collection, compared with a quarter of the 2.1 billion people in Central and Southern Asia, but only 12 percent in Northern Africa and Western Asia, and just 3 percent in Latin America and the Caribbean (WHO/UNICEF, 2023). For sanitation, over 3 hours were saved per week in South and Western Asia and in sub-Saharan Africa. Most studies did not disaggregate by respondent sex or age, with ten studies (out of 41 included studies) disaggregating findings by sex, seven studies including children, and five studies examining solely women, children or vulnerable groups. Of studies disaggregating by sex, the meta-analysis suggested large and significant effects of water supply interventions for women (d=- 0.24, 95%CI=-0.46, -0.01; 5 estimates), and no significant intervention effects for men (Figure 7). Only two studies were of the effects of programmes that specifically aimed to enhance access to water and sanitation services by vulnerable groups including people with disabilities (54,75); no studies of regular WASH examined effects by disability subgroups (including chronically ill and older adults), which is an important gap as these groups experience very large reductions in travel and access time (Table A6). No studies examined groups without fixed households (e.g. people with insecure housing) or who spend significant amounts of time away from their household (e.g. pastoralists), nor gender minorities. As time savings on WASH access may be particularly important for certain vulnerable subgroups, there is a need for further research collecting and presenting disaggregated data, even where the studies do not find statistically significant differences in effects.

### 5.1 Gender implications for WASH access and time reallocation

Globally, in 63 percent of households where water collection is conducted (16% of the population), women are primarily responsible for water carriage, compared with 26 percent where men are responsible (WHO/UNICEF, 2023). Similarly, in our systematic review and meta-analysis we found large and significant effects of water supply interventions for women but no significant intervention effects overall were found for men. While we did not look at water collection transport, men are more likely to collect water with time-efficient technology such as bikes, motorcycles or vehicles (80).

Reducing travel time for water collection offers potential to address gender inequalities if time savings are reallocated in a way that has positive benefits. In settings where women or girls have limited control over their households’ roles and agency due to gender norms and roles, travel time reductions may result in loss of mobility or be reallocated to other unpaid drudgery, and thus it is important to assess these wider changes when evaluating impacts. In the case of girls, the studies found medium-sized effects of water supply interventions on time reallocated to school, but no significant effects for boys. However, among adults, the meta-analysis showed no significant differences in time use for other activities, such as work or leisure, albeit over a very small sample of studies and contexts (Figure 10, Table 6). This indicates a clear need for further research to understand how time is reallocated to paid or unpaid work as well as other activities, potentially requiring improved measurement methods. Such evidence could highlight the role of WASH in contributing to the “3Rs” goal to recognise, reduce and redistribute to address the gendered burden of unpaid work such as water collection, as part of achieving SDG 5.4. Aside from reallocation of time, reduced time collection water can mean improved physical health due to decreased risk of injury and assault associated with water collection (4).

### 5.2 Gaps in evidence on sanitation and hygiene

Our findings provide evidence supporting the often-assumed benefit of sanitation interventions, reducing access time by 3.5 hours per week. Travelling to a safe place for open defecation or a shared sanitation facility, as well as time spent queueing at certain moments of the day, more adversely impacts women, including risks sexual violence and loss of privacy and dignity(81). No studies were identified that measured time allocation related to access to or use of hygiene facilities (e.g. hand-washing stations, shower blocks, or menstrual waste receptacles), indicating a need for research in this area, especially with growing calls for higher levels of household hygiene needed to prevent childhood ill-health (82) and mortality (Sharma Waddington et al., 2023).

### 5.3 Methods and research gaps relating to travel time and time use measurement

Our results suggest more research is needed on methods for measuring time use as most studies used self-reported surveys to measure time outcome, with the exceptions of Almanzar (2017) using time diaries to collect data on time use, Cairncross and Cliff (1987) observing women’s behaviour over the course of the waking day, and Winter et al. (2021) incorporating GPS for a subsample. Self-reports can be prone to bias for a range of reasons including recall bias, and depending on the context it may be important to have more accurate and detailed information to best fit different interventions or monitoring activities (83). Our findings also highlight the need for standardized questionnaires, as different time scales were used in different studies, mostly using time per trip or hours per week, which limited comparison and synthesis of results.

Furthermore, most studies in this review used fairly coarse measures of time use, without disaggregating by activity, such as travel to water sources or payment locations, waiting in line, household water management such as conducting treatment, cleaning storage containers, or time wasted due to unreliable piped water sources or waiting for water deliveries. Information on factors that slow down or speed up travel access time would also provide finer evidence, such as if vehicles were used or whether a longer route was taken due to safety (due to risks of road traffic, animals or assaults), or lack of water rights and abuse from landowners (84). More disaggregation can highlight time uses that may be overlooked and could be targeted by novel interventions.

In addition, data are needed, not just on quantity, but on quality and preferences for time use. Regarding quality, studies generally do not provide information on secondary activities, such as whether social activities were also conducted, whether young children are brought to water or defaecation sites when they are used, or whether another family members like an older girl is required to care for young children at these times. This is important, as such activities can increase the actual time or energy taken for the task (83). Advances used by researchers in other sectors such as agriculture offer potential methodological innovations for the water sector. For example,

Srinivasan et al., (2020) integrated data on time use with data on energy expenditure using accelerometers worn by study participants. Similarly, it is important to understand people’s preferences and bargaining power when time savings are reallocated for using that time. In this vein, Sinharoy et al., (2023) propose a measure of time agency that assesses choice over the allocation of one’s time. Finally, time is also closely related to convenience and therefore use of services; interventions that increase the burden of time, even by a small amount, may not be favoured under time budget constraints, particularly if the benefits are also not clearly observed (86).

### 5.4 Strengths and limitations

We conducted a systematic review and meta-analysis that combined studies from two recent systematic maps, done collaboratively. We searched for academic studies and grey literature, as well as studies in languages other than English (Spanish). We also aimed to make best use of existing evidence, by including uncontrolled studies with measurement before and after a WASH intervention, which are usually excluded from systematic reviews on the grounds of confounding. We also synthesised a range of outcome quantities, including time measured in natural units of minutes per trip and hours per week, and using standardised mean difference (*d*) effect sizes, which are calculable from information commonly reported in studies such as t-statistics. However, the study has several limitations. As we are synthesising evidence across a range of interventions and counterfactual WASH service starting points, there is imprecision in the relationships being measured. There are also limitations to generalisability of the evidence, since some interventions like community connections were only evaluated in some parts of the world such as sub-Saharan

Africa. Perhaps the biggest limitation of this review is that we excluded any purely qualitative information from qualitative, quantitative or mixed-methods studies. Qualitative evidence would contribute to more comprehensively understanding WASH implementation contexts and the implications of the effects on time use for important factors like convenience. It would also help elucidate complex mechanisms underpinning some of the effects, particularly those relating to time use reallocation, which may help explain why generalisable effects were not apparent across the different contexts studied.

## 5. Conclusion

This study provides a systematic review and meta-analysis of time use for accessing WASH services and time reallocation to other activities, following WASH interventions in L&MIC contexts. Our results showing significant effects of water supply and sanitation interventions on time savings, including particularly for women. This provides evidence in support of efforts to reach SDG target 6.1 for drinking water provided on premises and SDG target 6.2 for household-level latrines, as well as for better collecting and tracking of this information (87). We also found significant effects of WASH interventions for girls’ schooling, providing evidence to support SDG targets 4.1 on school enrolment and attainment, and 4.5 on eliminating educational disparities linked to gender and vulnerable status. While WASH travel and access time savings are often purported by civil society organizations to be allocated to income-generating activities or health-seeking behaviours and thus benefitting women, we found inadequate evidence on ways that time was reallocated, except in the case of girls’ reallocation of time to education.

There is a clear need for more research in this area. Even a small increase in time to access WASH amenities, as we found for water quality interventions (and may well have found for hygiene interventions, if any studies had measured it), is likely to be associated with a reduction in convenience for those already experiencing time poverty, and therefore a reduction in demand. It follows that better integrated concepts of time and time measurement in WASH practice and research may help in the design of interventions that are more effective in achieving desired goals of behaviour change and improvements in health and quality of life. This synthesis contributes to the growing evidence base on evaluating the wider societal and health benefits of WASH interventions, beyond child diarrheal disease (88), providing impetus for efforts to prioritize the needed resources to achieve universal WASH coverage. In particular, the clear effects for women and girls indicates potential contributions to women’s and girl’s health and gender equality by reducing the load of unpaid water collection work and the burden of travel time for safe sanitation.

## Data Availability

All data produced in the present work are contained in the manuscript

## Availability of data, code and other materials

All data used for this review are included in the manuscript.

## Supplementary information

### Annex 1: Measures of effect

The standardised mean difference (*d*) measures the size of the intervention effect in each study in units of standard deviation observed in that study and is thus independent of units of measurement. The *d* statistic is the ratio of the mean difference, where yt is the outcome in the WASH intervention group and yc the outcome in the comparison group, to the standard deviation of the outcome, S(y):

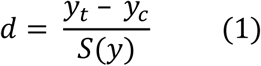

For the denominator, S(y), the pooled standard deviation 𝑆_𝑝_ was calculated:

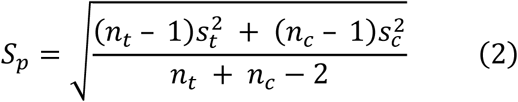

The 95 percent confidence intervals used the standard error of *d*, se(d), given by:

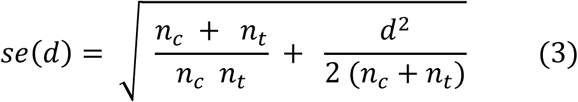

In cases where outcomes were reported in frequencies, such as households whose travel times were less than or greater than 30 minutes, the Cox-transformed log odds ratio (OR) was estimated:

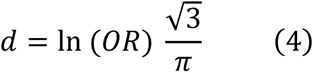

The Standard error of Cox-transformed *d* is given as:

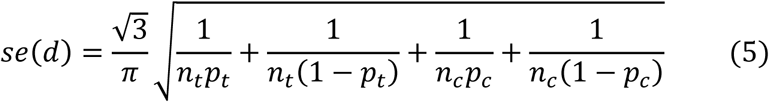

For studies reporting effect sizes from regression estimates on outcomes, then:

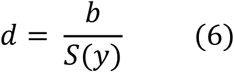

where b is the (mean difference) coefficient estimated in the regression. Where regression studies did not report S(y), the standard error se(b) of the test statistic for effect size estimate b was usually available or could be calculated. In such cases, the pooled standard deviation was calculated using (Lipsey and Wilson, 2021):

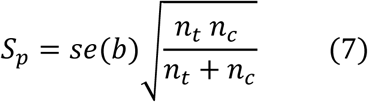

### Annex 2 Overview of included studies

Table A1 describes drinking water interventions while Table A2 describe sanitation interventions. These tables report details about each including country focus, intervention details, study design, outcomes measured, targeted participant group and baselines. Overall, the risk-of-bias assessment found that two studies (5%) were rated as at ‘low risk’ in attributing the change in time to the WASH intervention (Biran et al., 2018; Winter et al., 2021), 11 studies (27%) were rated at ‘moderate risk’ and the remaining studies (n=28, 68%) were rated at ‘high risk of bias’. Most studies were at ‘low risk of bias’ due to confounding, partly because travel time is subject to low risk of confounding anyway, so even an uncontrolled before versus after (BA) study can produce an unbiased estimate of travel time provided it is measured shortly after implementation of the WASH improvement. Reporting bias was potentially more problematic, however, especially where studies used recall of baseline measures. One study with 5-year follow-up noted that "other factors external to the water project, such as the development of a new road infrastructure in some of the sampled communities could have impacted the level of participation by people in activities that required travel by road to such places as markets within and outside the area" (Arku, 2010: p.236). Most studies (93%) used self-report surveys to measure travel time and time use reallocation, with the exceptions of Almanzar et al. (2017) who used time diaries, Cairncross et al. (1987) who observed women’s behaviour over the course of the waking day, and Winter et al. (2021) who used GPS for a subsample of observations and otherwise self-report.

**Table A1.**
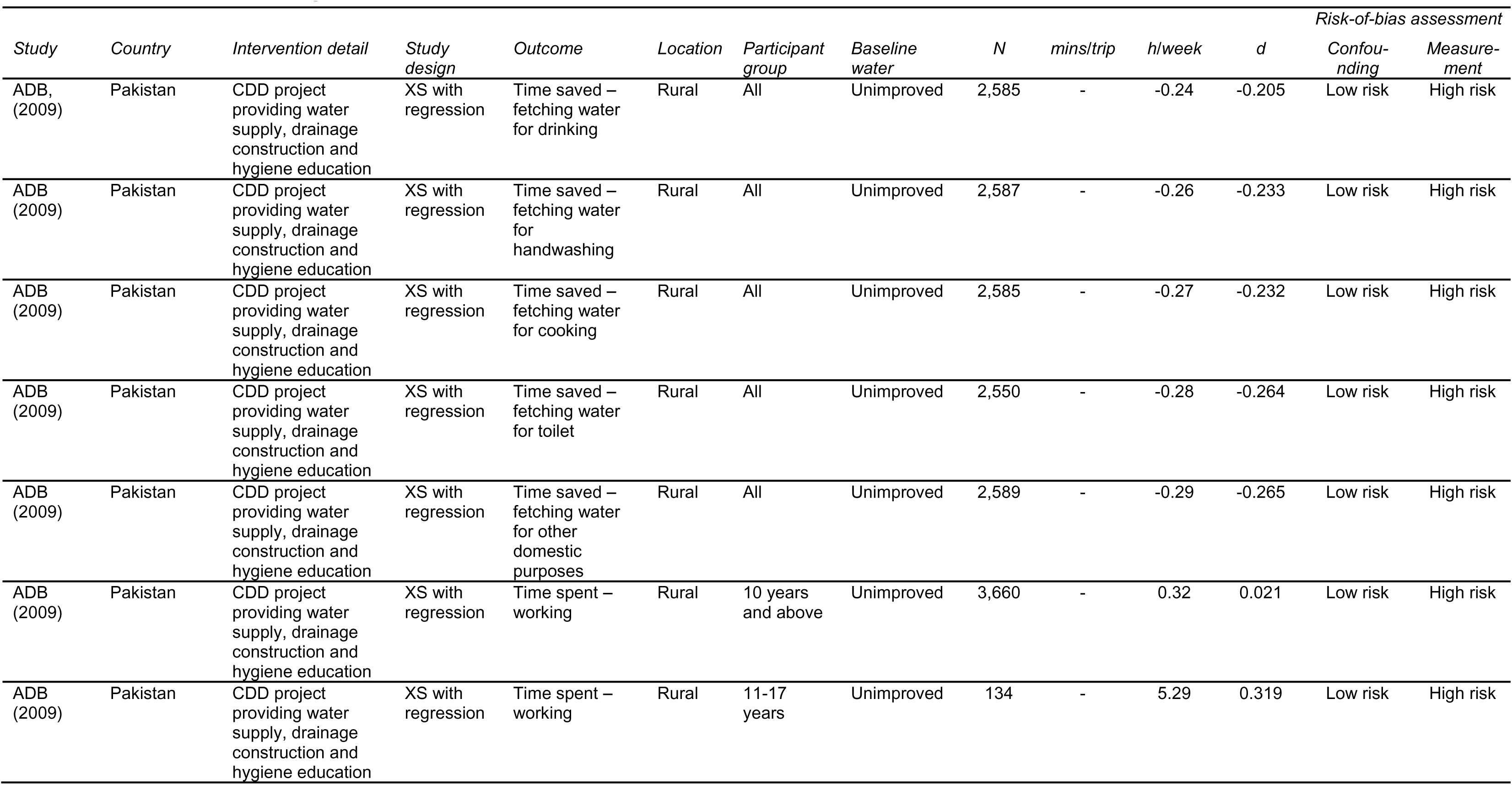
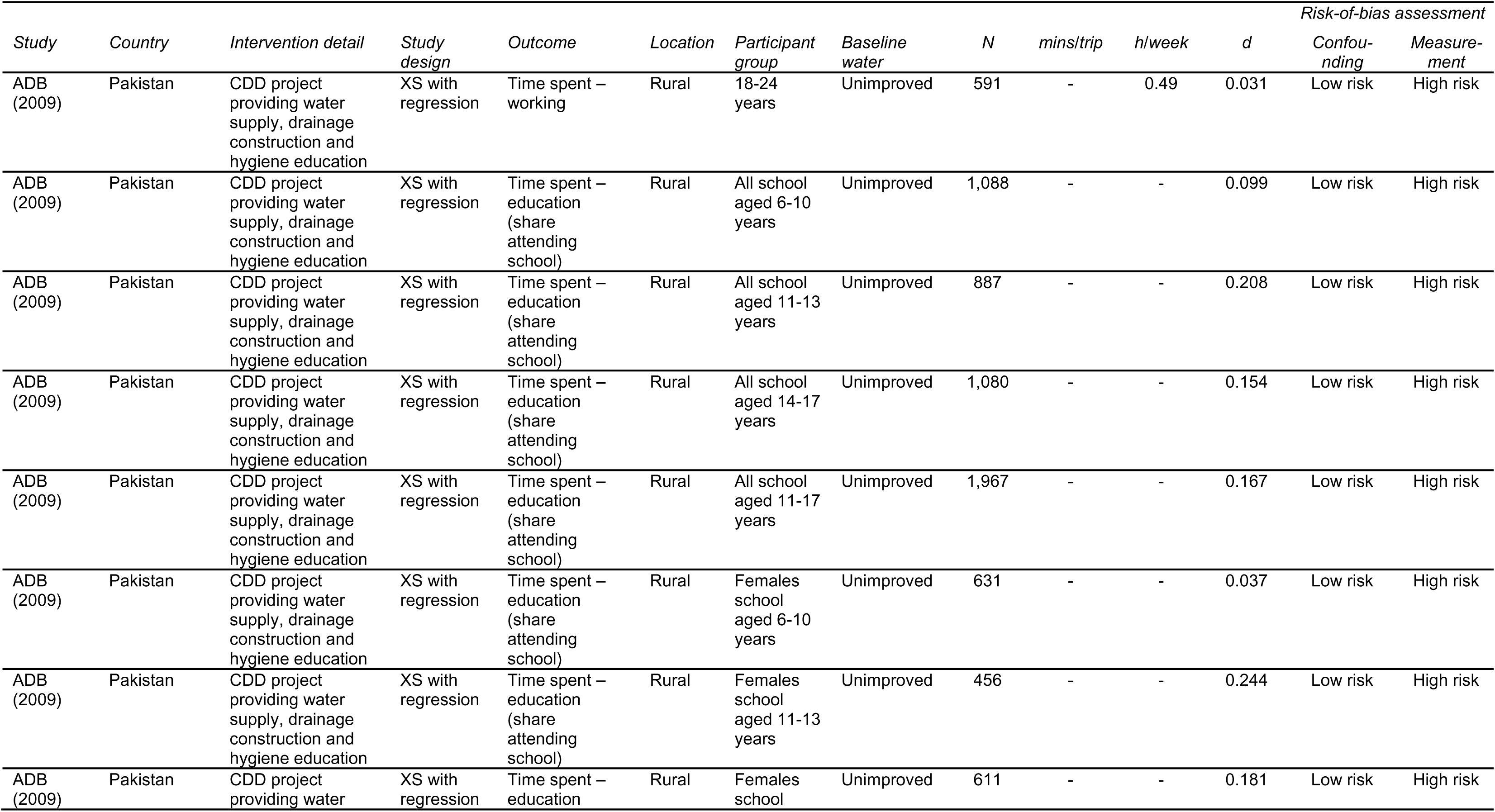
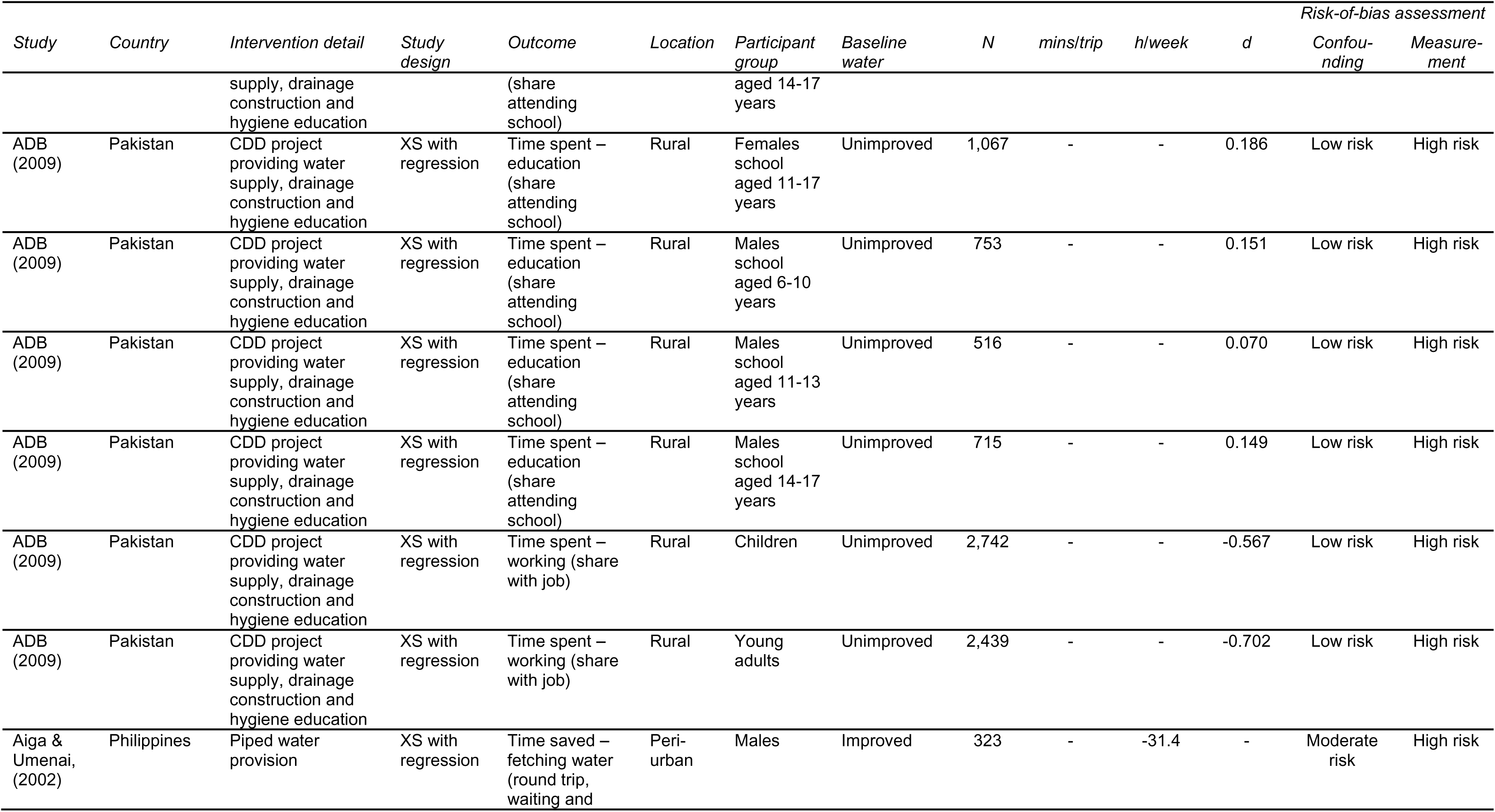
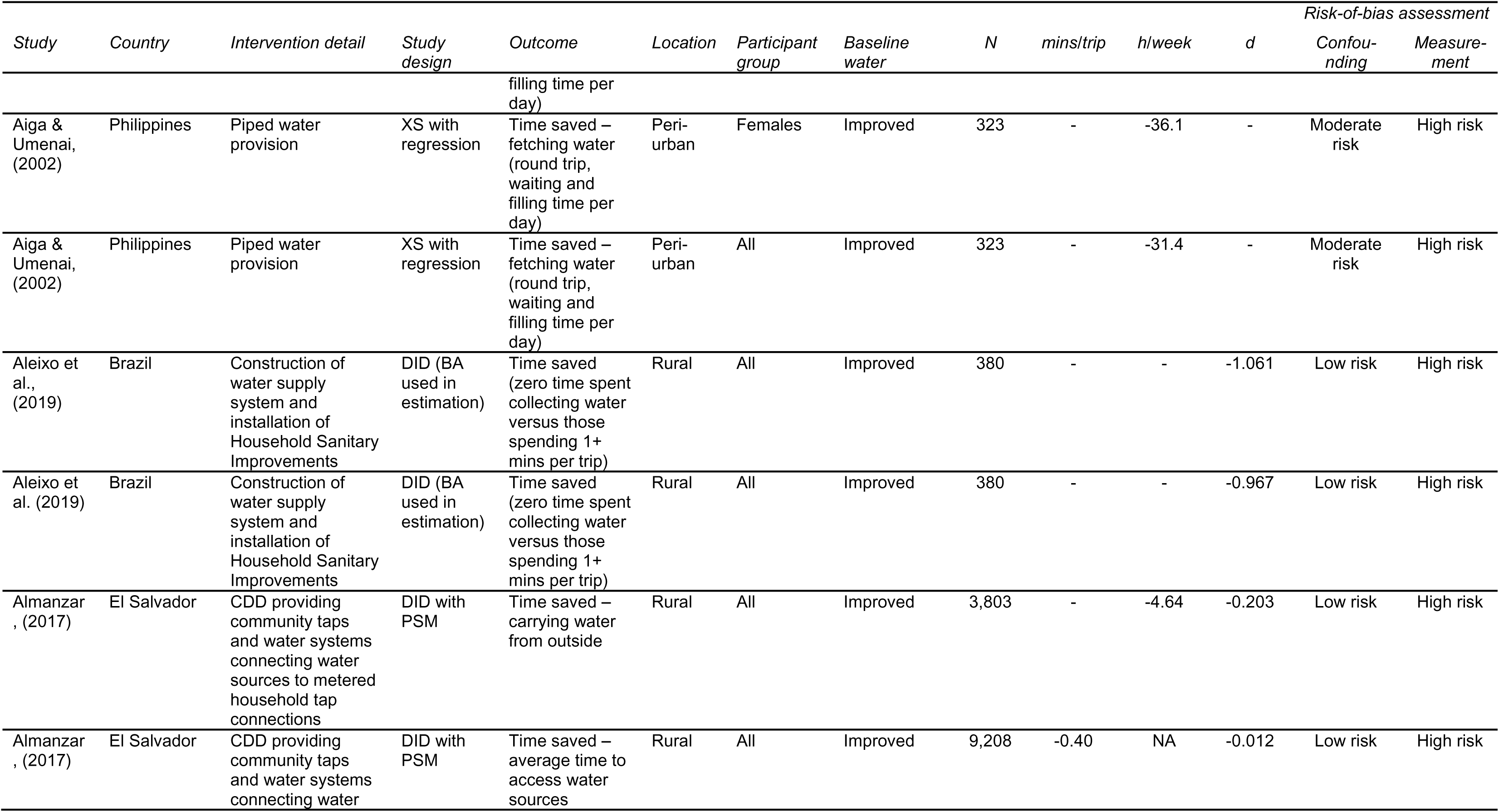
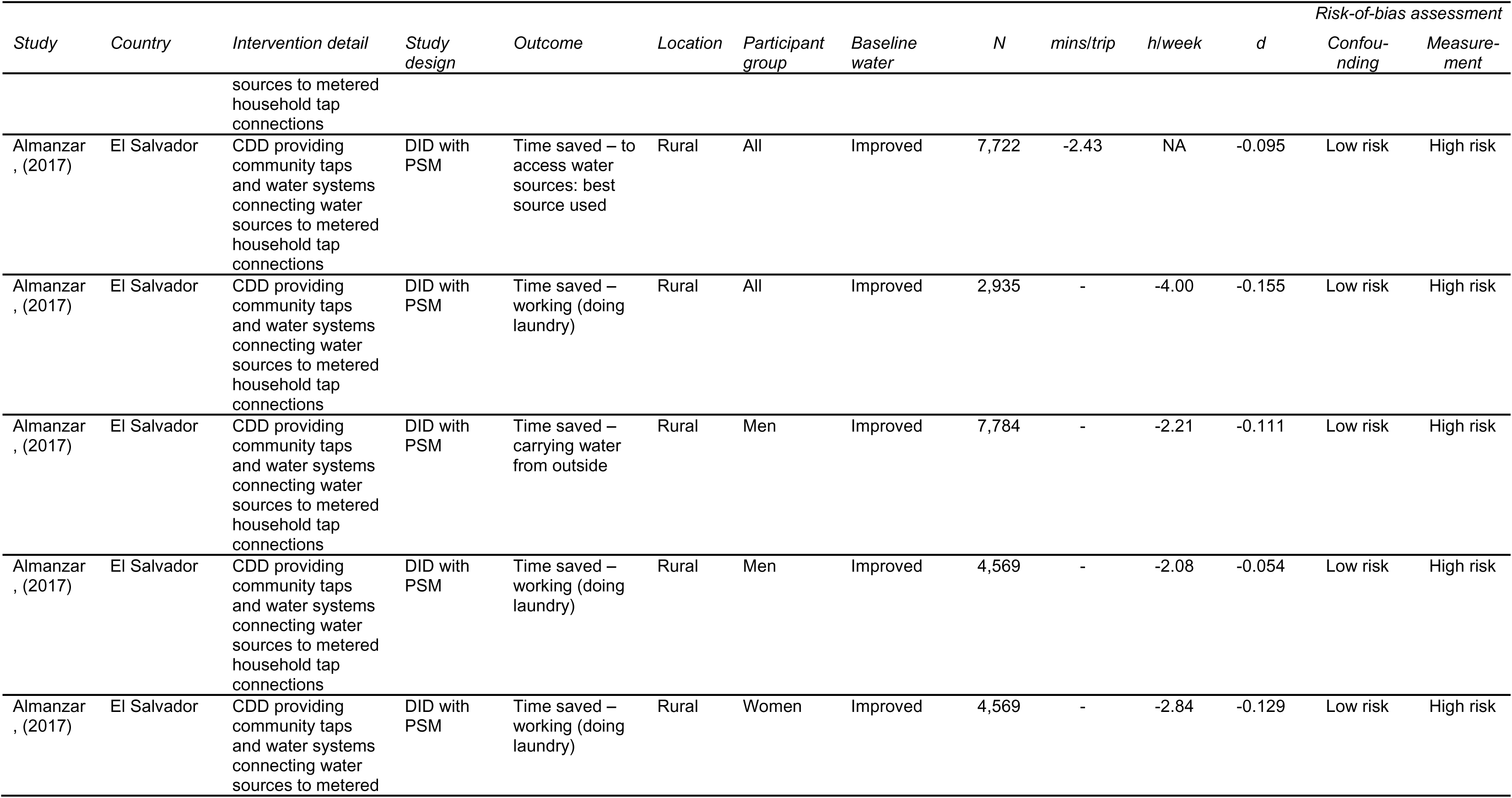
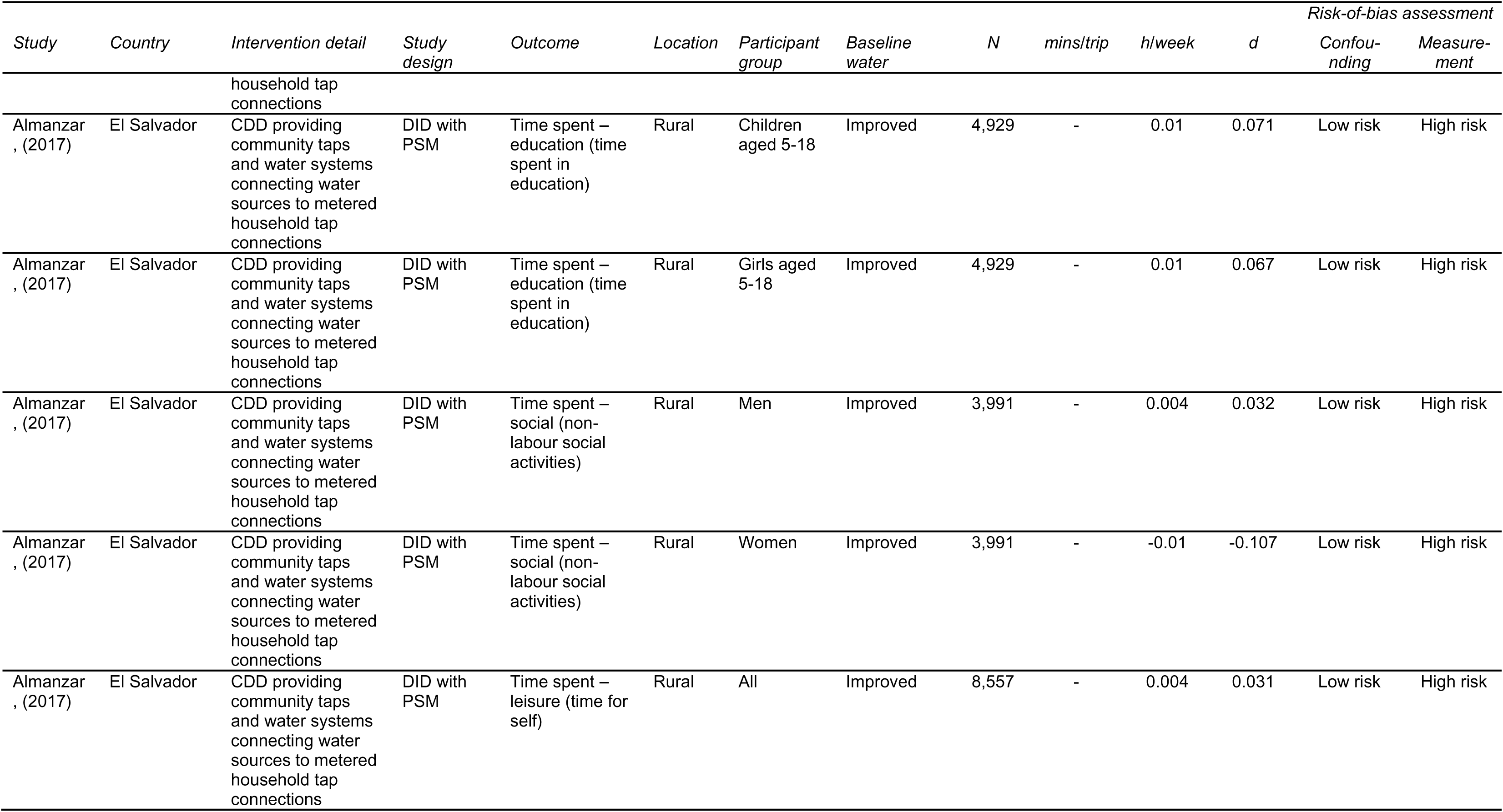
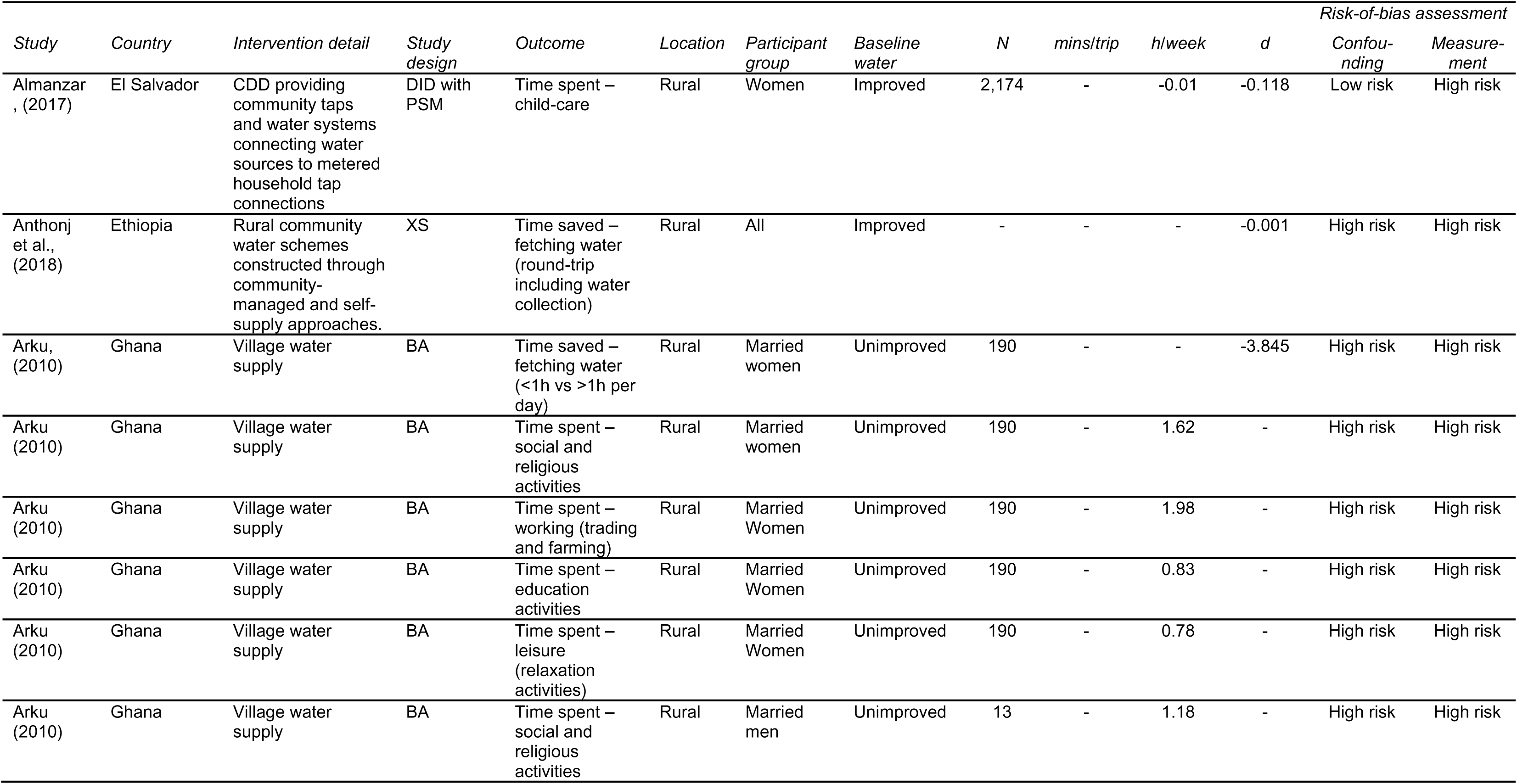
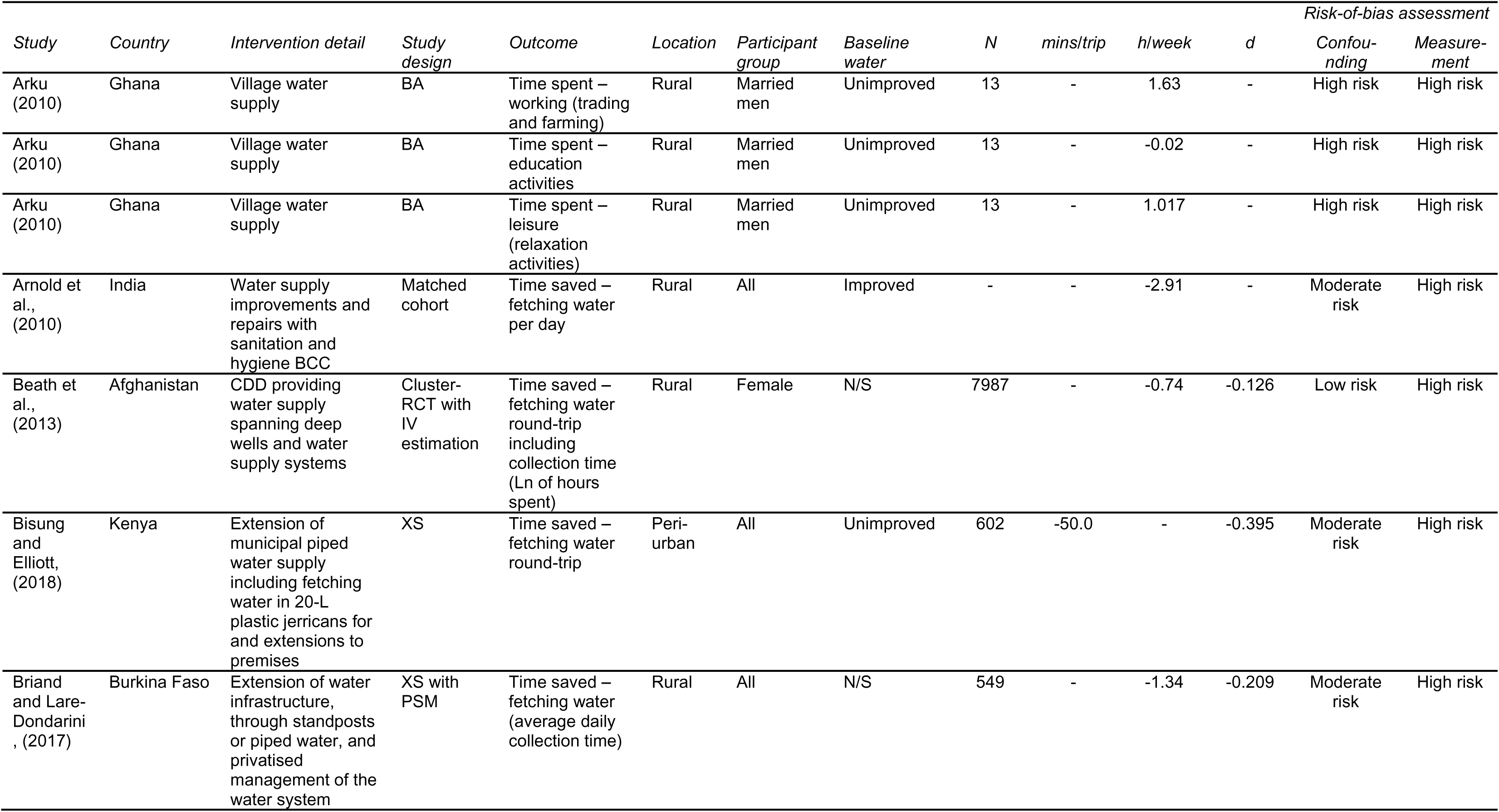
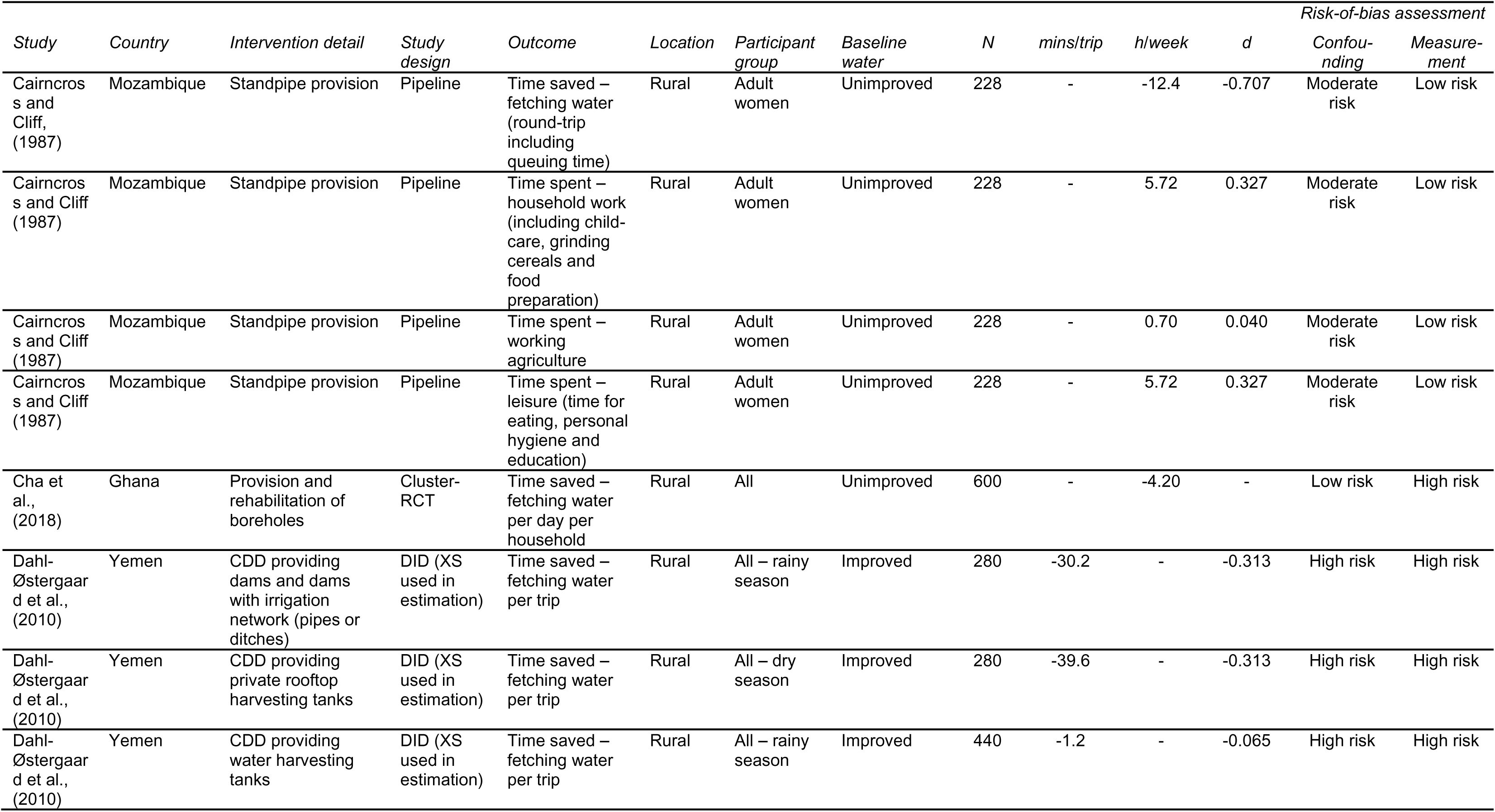
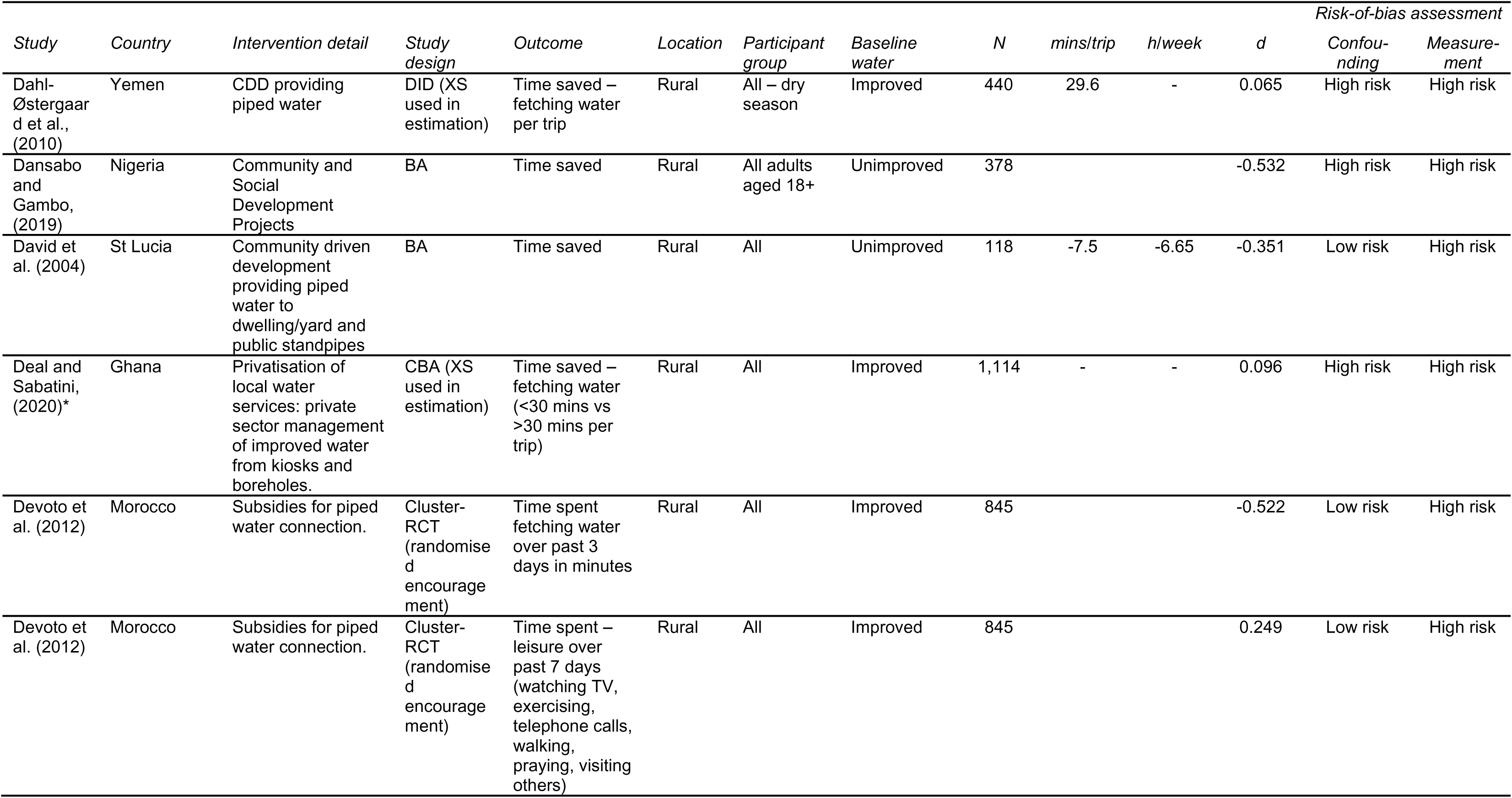
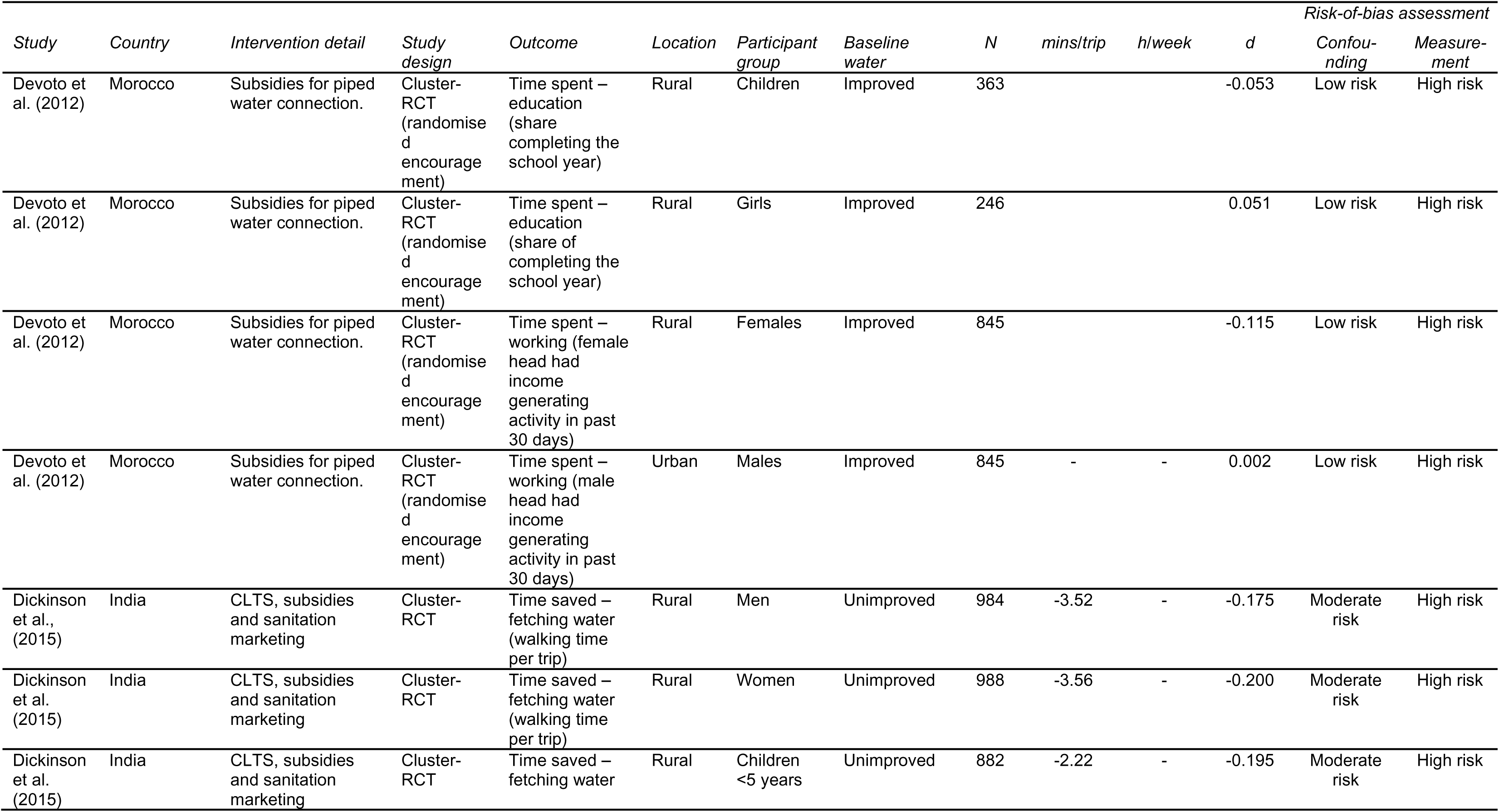
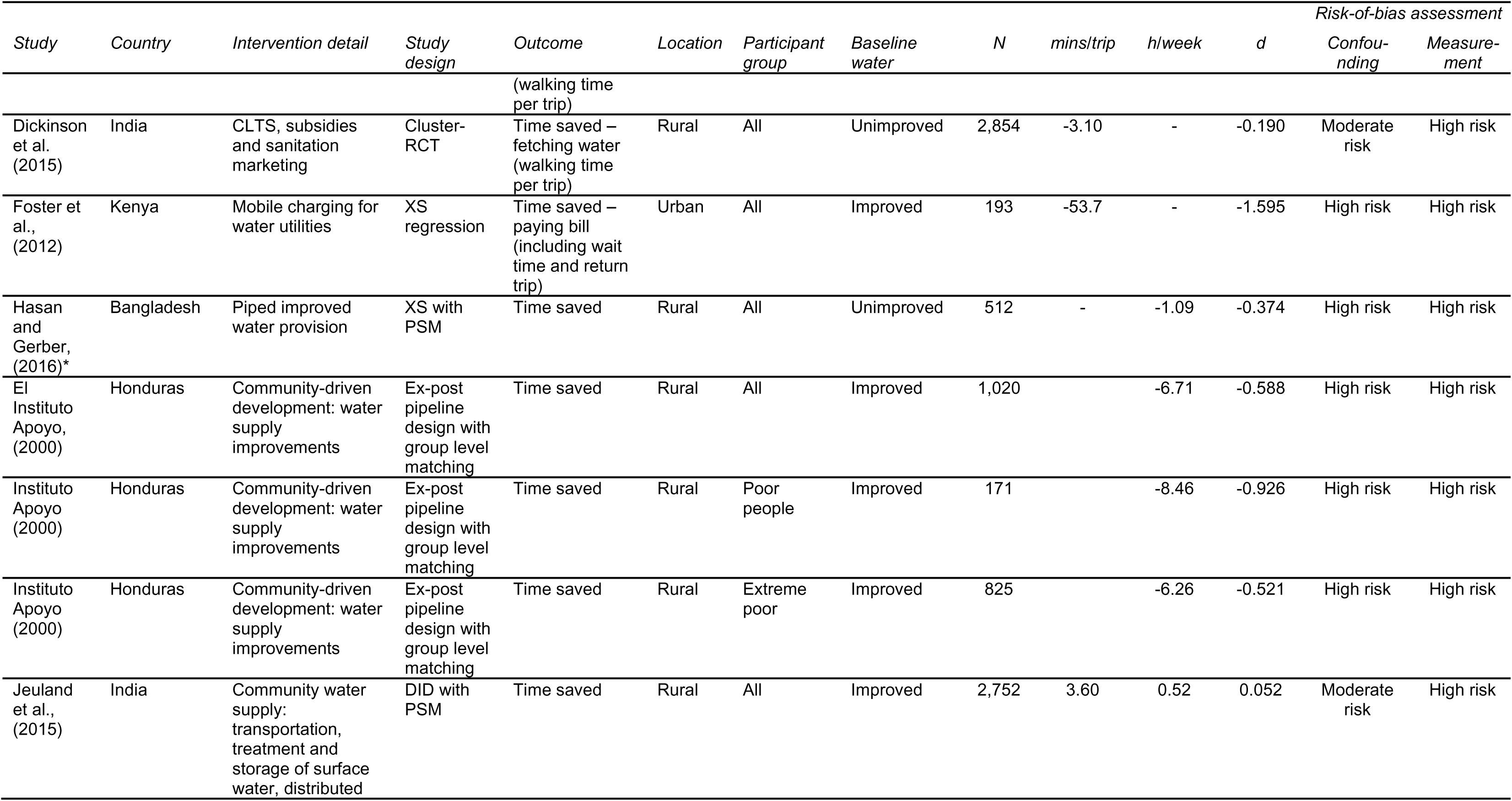
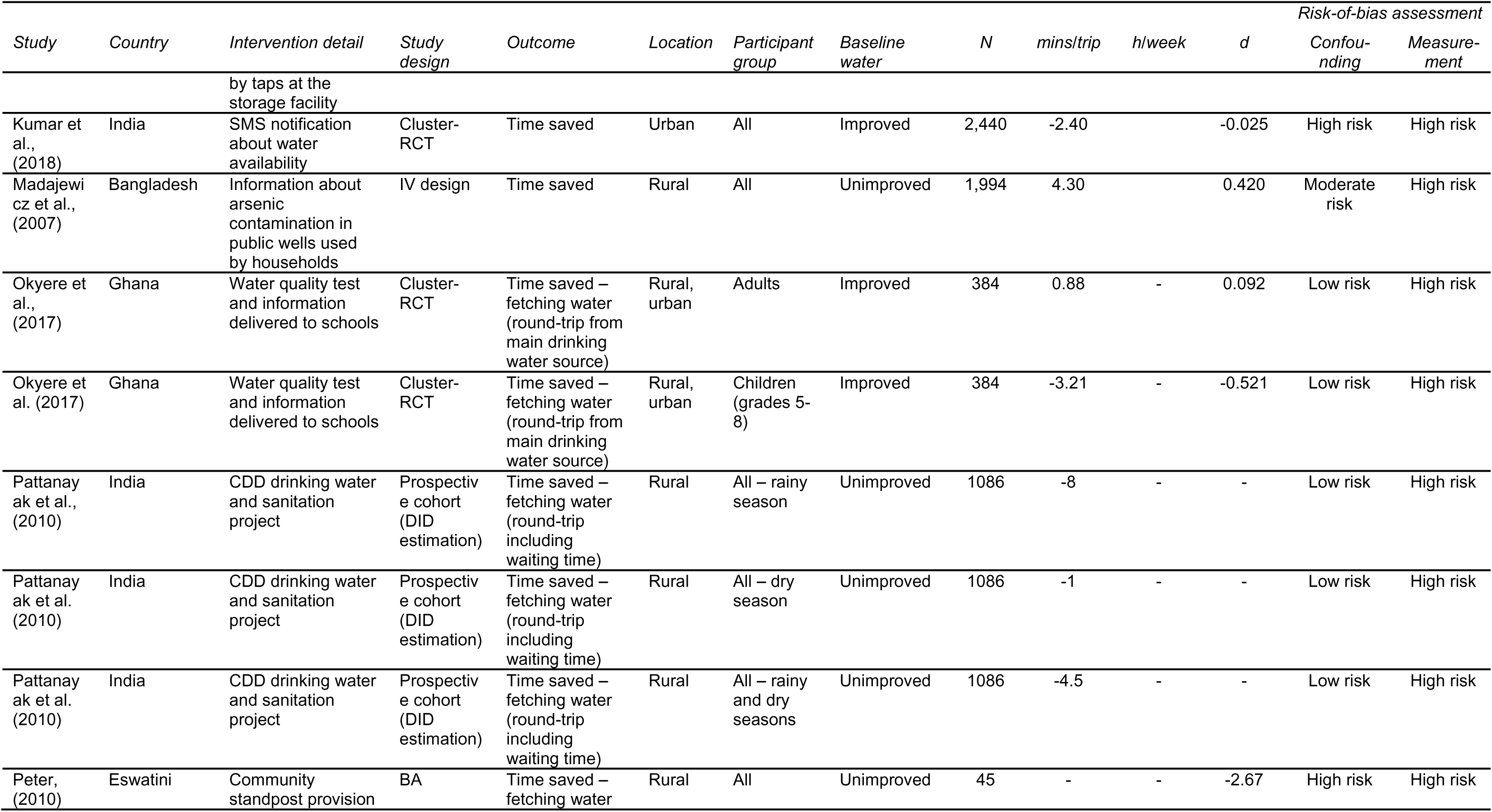
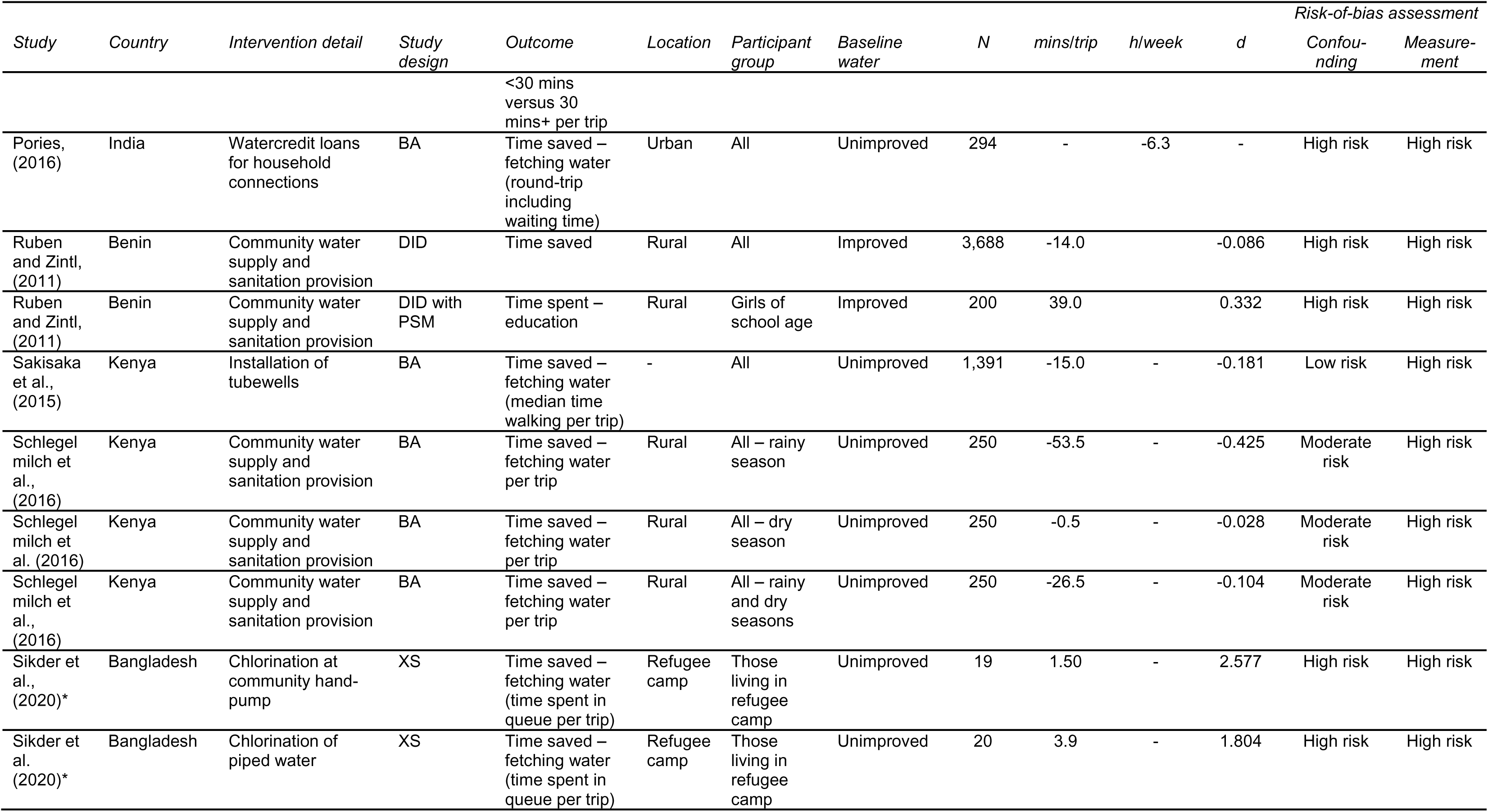
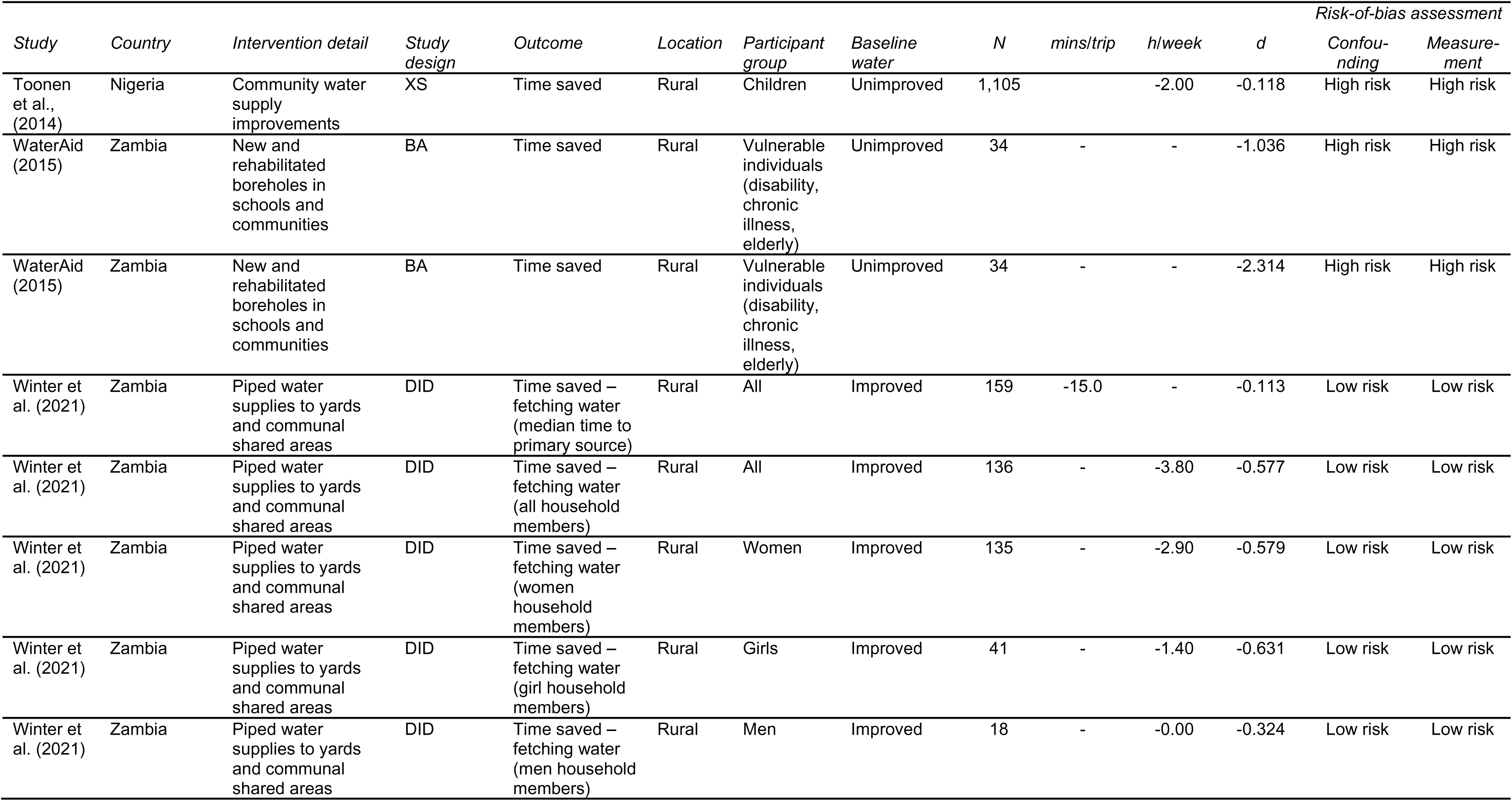
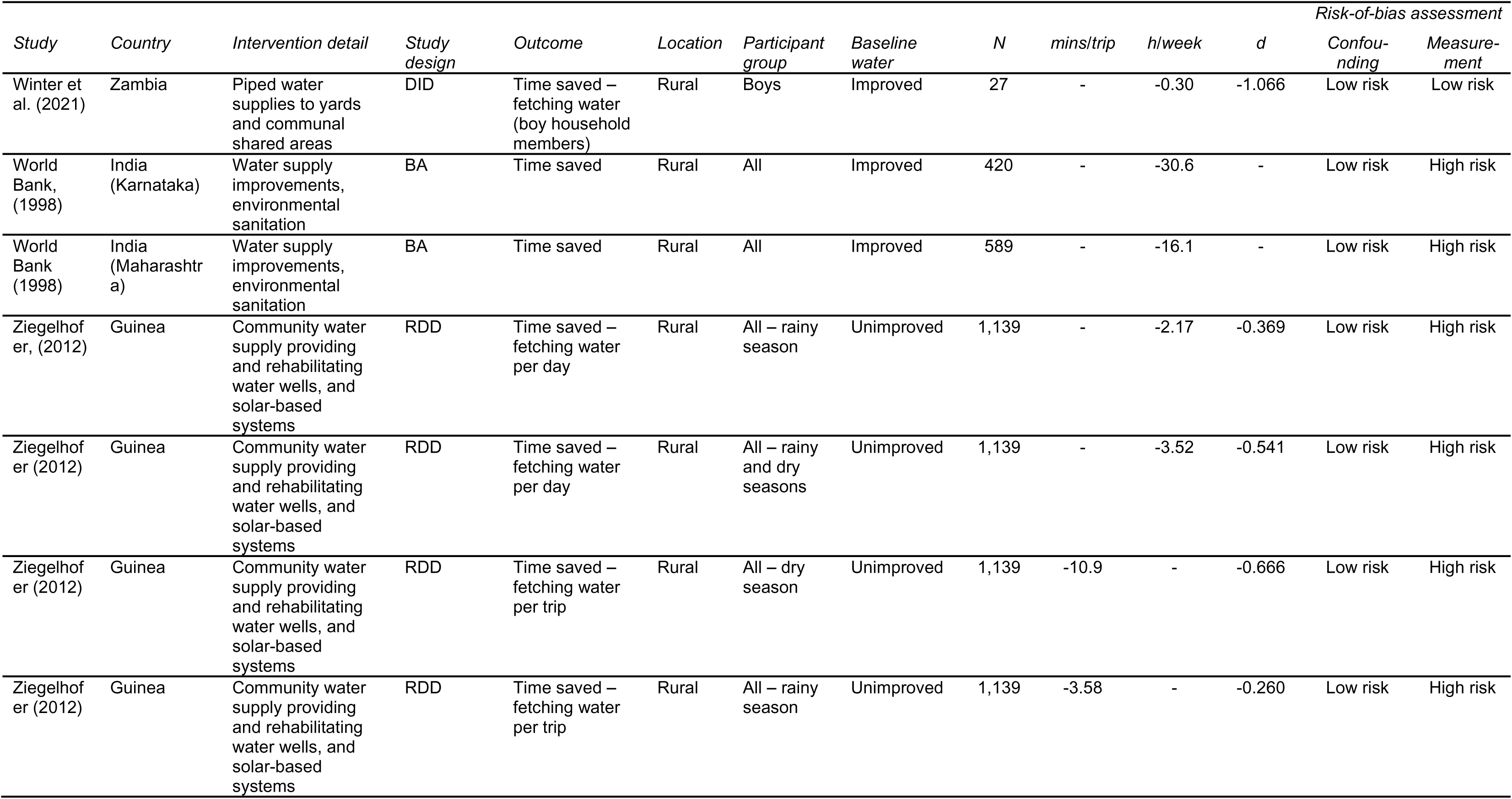
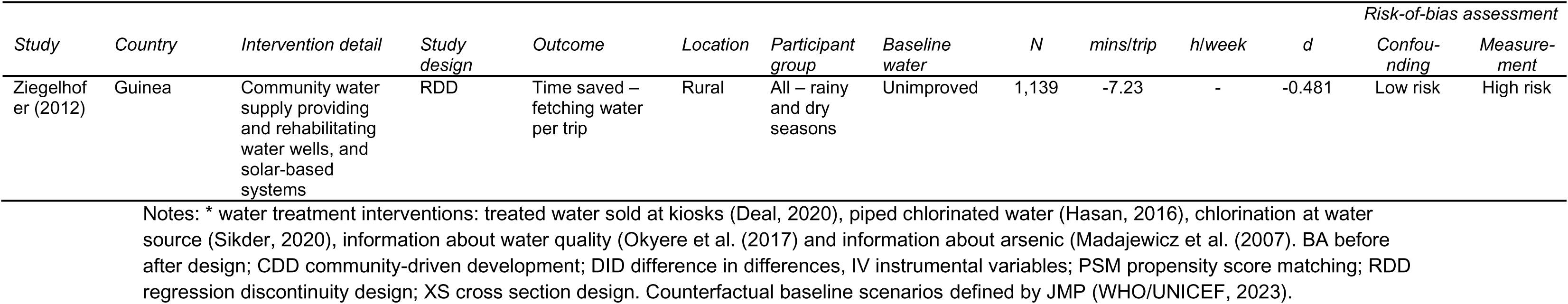
Drinking water interventions.

| Study | Country | Intervention detail | Study design | Outcome | Location | Participant group | Baseline water | N | mins/trip | h/week | d | Risk-of-bias assessment |  |
| --- | --- | --- | --- | --- | --- | --- | --- | --- | --- | --- | --- | --- | --- |
|  |  |  |  |  |  |  |  |  |  |  |  | Confounding | Measurement |
| ADB, (2009) | Pakistan | CDD project providing water supply, drainage construction and hygiene education | XS with regression | Time saved – fetching water for drinking | Rural | All | Unimproved | 2,585 | - | -0.24 | -0.205 | Low risk | High risk |
| ADB (2009) | Pakistan | CDD project providing water supply, drainage construction and hygiene education | XS with regression | Time saved – fetching water for handwashing | Rural | All | Unimproved | 2,587 | - | -0.26 | -0.233 | Low risk | High risk |
| ADB (2009) | Pakistan | CDD project providing water supply, drainage construction and hygiene education | XS with regression | Time saved – fetching water for cooking | Rural | All | Unimproved | 2,585 | - | -0.27 | -0.232 | Low risk | High risk |
| ADB (2009) | Pakistan | CDD project providing water supply, drainage construction and hygiene education | XS with regression | Time saved – fetching water for toilet | Rural | All | Unimproved | 2,550 | - | -0.28 | -0.264 | Low risk | High risk |
| ADB (2009) | Pakistan | CDD project providing water supply, drainage construction and hygiene education | XS with regression | Time saved – fetching water for other domestic purposes | Rural | All | Unimproved | 2,589 | - | -0.29 | -0.265 | Low risk | High risk |
| ADB (2009) | Pakistan | CDD project providing water supply, drainage construction and hygiene education | XS with regression | Time spent – working | Rural | 10 years and above | Unimproved | 3,660 | - | 0.32 | 0.021 | Low risk | High risk |
| ADB (2009) | Pakistan | CDD project providing water supply, drainage construction and hygiene education | XS with regression | Time spent – working | Rural | 11-17 years | Unimproved | 134 | - | 5.29 | 0.319 | Low risk | High risk |
|  |  |  |  |  |  |  |  |  |  |  |  | Confounding | Measurement |
| ADB (2009) | Pakistan | CDD project providing water supply, drainage construction and hygiene education | XS with regression | Time spent – working | Rural | 18-24 years | Unimproved | 591 | - | 0.49 | 0.031 | Low risk | High risk |
| ADB (2009) | Pakistan | CDD project providing water supply, drainage construction and hygiene education | XS with regression | Time spent – education (share attending school) | Rural | All school aged 6-10 years | Unimproved | 1,088 | - | - | 0.099 | Low risk | High risk |
| ADB (2009) | Pakistan | CDD project providing water supply, drainage construction and hygiene education | XS with regression | Time spent – education (share attending school) | Rural | All school aged 11-13 years | Unimproved | 887 | - | - | 0.208 | Low risk | High risk |
| ADB (2009) | Pakistan | CDD project providing water supply, drainage construction and hygiene education | XS with regression | Time spent – education (share attending school) | Rural | All school aged 14-17 years | Unimproved | 1,080 | - | - | 0.154 | Low risk | High risk |
| ADB (2009) | Pakistan | CDD project providing water supply, drainage construction and hygiene education | XS with regression | Time spent – education (share attending school) | Rural | All school aged 11-17 years | Unimproved | 1,967 | - | - | 0.167 | Low risk | High risk |
| ADB (2009) | Pakistan | CDD project providing water supply, drainage construction and hygiene education | XS with regression | Time spent – education (share attending school) | Rural | Females school aged 6-10 years | Unimproved | 631 | - | - | 0.037 | Low risk | High risk |
| ADB (2009) | Pakistan | CDD project providing water supply, drainage construction and hygiene education | XS with regression | Time spent – education (share attending school) | Rural | Females school aged 11-13 years | Unimproved | 456 | - | - | 0.244 | Low risk | High risk |
| ADB (2009) | Pakistan | CDD project providing water | XS with regression | Time spent – education | Rural | Females school | Unimproved | 611 | - | - | 0.181 | Low risk | High risk |
|  |  |  |  |  |  |  |  |  |  |  |  | Confounding | Measurement |
|  |  | supply, drainage construction and hygiene education |  | (share attending school) |  | aged 14-17 years |  |  |  |  |  |  |  |
| ADB (2009) | Pakistan | CDD project providing water supply, drainage construction and hygiene education | XS with regression | Time spent – education (share attending school) | Rural | Females school aged 11-17 years | Unimproved | 1,067 | - | - | 0.186 | Low risk | High risk |
| ADB (2009) | Pakistan | CDD project providing water supply, drainage construction and hygiene education | XS with regression | Time spent – education (share attending school) | Rural | Males school aged 6-10 years | Unimproved | 753 | - | - | 0.151 | Low risk | High risk |
| ADB (2009) | Pakistan | CDD project providing water supply, drainage construction and hygiene education | XS with regression | Time spent – education (share attending school) | Rural | Males school aged 11-13 years | Unimproved | 516 | - | - | 0.070 | Low risk | High risk |
| ADB (2009) | Pakistan | CDD project providing water supply, drainage construction and hygiene education | XS with regression | Time spent – education (share attending school) | Rural | Males school aged 14-17 years | Unimproved | 715 | - | - | 0.149 | Low risk | High risk |
| ADB (2009) | Pakistan | CDD project providing water supply, drainage construction and hygiene education | XS with regression | Time spent – working (share with job) | Rural | Children | Unimproved | 2,742 | - | - | -0.567 | Low risk | High risk |
| ADB (2009) | Pakistan | CDD project providing water supply, drainage construction and hygiene education | XS with regression | Time spent – working (share with job) | Rural | Young adults | Unimproved | 2,439 | - | - | -0.702 | Low risk | High risk |
| Aiga & Umenai, (2002) | Philippines | Piped water provision | XS with regression | Time saved – fetching water (round trip, waiting and | Peri-urban | Males | Improved | 323 | - | -31.4 | - | Moderate risk | High risk |
|  |  |  |  |  |  |  |  |  |  |  |  | Confounding | Measurement |
|  |  |  |  | filling time per day) |  |  |  |  |  |  |  |  |  |
| Aiga & Umenai, (2002) | Philippines | Piped water provision | XS with regression | Time saved – fetching water (round trip, waiting and filling time per day) | Peri-urban | Females | Improved | 323 | - | -36.1 | - | Moderate risk | High risk |
| Aiga & Umenai, (2002) | Philippines | Piped water provision | XS with regression | Time saved – fetching water (round trip, waiting and filling time per day) | Peri-urban | All | Improved | 323 | - | -31.4 | - | Moderate risk | High risk |
| Aleixo et al., (2019) | Brazil | Construction of water supply system and installation of Household Sanitary Improvements | DID (BA used in estimation) | Time saved (zero time spent collecting water versus those spending 1+ mins per trip) | Rural | All | Improved | 380 | - | - | -1.061 | Low risk | High risk |
| Aleixo et al. (2019) | Brazil | Construction of water supply system and installation of Household Sanitary Improvements | DID (BA used in estimation) | Time saved (zero time spent collecting water versus those spending 1+ mins per trip) | Rural | All | Improved | 380 | - | - | -0.967 | Low risk | High risk |
| Almanzar , (2017) | El Salvador | CDD providing community taps and water systems connecting water sources to metered household tap connections | DID with PSM | Time saved – carrying water from outside | Rural | All | Improved | 3,803 | - | -4.64 | -0.203 | Low risk | High risk |
| Almanzar , (2017) | El Salvador | CDD providing community taps and water systems connecting water | DID with PSM | Time saved – average time to access water sources | Rural | All | Improved | 9,208 | -0.40 | NA | -0.012 | Low risk | High risk |
|  |  |  |  |  |  |  |  |  |  |  |  | Confounding | Measurement |
|  |  | sources to metered household tap connections |  |  |  |  |  |  |  |  |  |  |  |
| Almanzar , (2017) | El Salvador | CDD providing community taps and water systems connecting water sources to metered household tap connections | DID with PSM | Time saved – to access water sources: best source used | Rural | All | Improved | 7,722 | -2.43 | NA | -0.095 | Low risk | High risk |
| Almanzar , (2017) | El Salvador | CDD providing community taps and water systems connecting water sources to metered household tap connections | DID with PSM | Time saved – working (doing laundry) | Rural | All | Improved | 2,935 | - | -4.00 | -0.155 | Low risk | High risk |
| Almanzar , (2017) | El Salvador | CDD providing community taps and water systems connecting water sources to metered household tap connections | DID with PSM | Time saved – carrying water from outside | Rural | Men | Improved | 7,784 | - | -2.21 | -0.111 | Low risk | High risk |
| Almanzar , (2017) | El Salvador | CDD providing community taps and water systems connecting water sources to metered household tap connections | DID with PSM | Time saved – working (doing laundry) | Rural | Men | Improved | 4,569 | - | -2.08 | -0.054 | Low risk | High risk |
| Almanzar , (2017) | El Salvador | CDD providing community taps and water systems connecting water sources to metered household tap connections | DID with PSM | Time saved – working (doing laundry) | Rural | Women | Improved | 4,569 | - | -2.84 | -0.129 | Low risk | High risk |
|  |  |  |  |  |  |  |  |  |  |  |  | Confounding | Measurement |
|  |  | household tap connections |  |  |  |  |  |  |  |  |  |  |  |
| Almanzar , (2017) | El Salvador | CDD providing community taps and water systems connecting water sources to metered household tap connections | DID with PSM | Time spent – education (time spent in education) | Rural | Children aged 5-18 | Improved | 4,929 | - | 0.01 | 0.071 | Low risk | High risk |
| Almanzar , (2017) | El Salvador | CDD providing community taps and water systems connecting water sources to metered household tap connections | DID with PSM | Time spent – education (time spent in education) | Rural | Girls aged 5-18 | Improved | 4,929 | - | 0.01 | 0.067 | Low risk | High risk |
| Almanzar , (2017) | El Salvador | CDD providing community taps and water systems connecting water sources to metered household tap connections | DID with PSM | Time spent – social (non-labour social activities) | Rural | Men | Improved | 3,991 | - | 0.004 | 0.032 | Low risk | High risk |
| Almanzar , (2017) | El Salvador | CDD providing community taps and water systems connecting water sources to metered household tap connections | DID with PSM | Time spent – social (non-labour social activities) | Rural | Women | Improved | 3,991 | - | -0.01 | -0.107 | Low risk | High risk |
| Almanzar , (2017) | El Salvador | CDD providing community taps and water systems connecting water sources to metered household tap connections | DID with PSM | Time spent – leisure (time for self) | Rural | All | Improved | 8,557 | - | 0.004 | 0.031 | Low risk | High risk |
|  |  |  |  |  |  |  |  |  |  |  |  | Confounding | Measurement |
| Almanzar , (2017) | El Salvador | CDD providing community taps and water systems connecting water sources to metered household tap connections | DID with PSM | Time spent – child-care | Rural | Women | Improved | 2,174 | - | -0.01 | -0.118 | Low risk | High risk |
| Anthonj et al., (2018) | Ethiopia | Rural community water schemes constructed through community-managed and self-supply approaches. | XS | Time saved – fetching water (round-trip including water collection) | Rural | All | Improved | - | - | - | -0.001 | High risk | High risk |
| Arku, (2010) | Ghana | Village water supply | BA | Time saved – fetching water (<1h vs >1h per day) | Rural | Married women | Unimproved | 190 | - | - | -3.845 | High risk | High risk |
| Arku (2010) | Ghana | Village water supply | BA | Time spent – social and religious activities | Rural | Married women | Unimproved | 190 | - | 1.62 | - | High risk | High risk |
| Arku (2010) | Ghana | Village water supply | BA | Time spent – working (trading and farming) | Rural | Married Women | Unimproved | 190 | - | 1.98 | - | High risk | High risk |
| Arku (2010) | Ghana | Village water supply | BA | Time spent – education activities | Rural | Married Women | Unimproved | 190 | - | 0.83 | - | High risk | High risk |
| Arku (2010) | Ghana | Village water supply | BA | Time spent – leisure (relaxation activities) | Rural | Married Women | Unimproved | 190 | - | 0.78 | - | High risk | High risk |
| Arku (2010) | Ghana | Village water supply | BA | Time spent – social and religious activities | Rural | Married men | Unimproved | 13 | - | 1.18 | - | High risk | High risk |
|  |  |  |  |  |  |  |  |  |  |  |  | Confounding | Measurement |
| Arku (2010) | Ghana | Village water supply | BA | Time spent – working (trading and farming) | Rural | Married men | Unimproved | 13 | - | 1.63 | - | High risk | High risk |
| Arku (2010) | Ghana | Village water supply | BA | Time spent – education activities | Rural | Married men | Unimproved | 13 | - | -0.02 | - | High risk | High risk |
| Arku (2010) | Ghana | Village water supply | BA | Time spent – leisure (relaxation activities) | Rural | Married men | Unimproved | 13 | - | 1.017 | - | High risk | High risk |
| Arnold et al., (2010) | India | Water supply improvements and repairs with sanitation and hygiene BCC | Matched cohort | Time saved – fetching water per day | Rural | All | Improved | - | - | -2.91 | - | Moderate risk | High risk |
| Beath et al., (2013) | Afghanistan | CDD providing water supply spanning deep wells and water supply systems | Cluster-RCT with IV estimation | Time saved – fetching water round-trip including collection time (Ln of hours spent) | Rural | Female | N/S | 7987 | - | -0.74 | -0.126 | Low risk | High risk |
| Bisung and Elliott, (2018) | Kenya | Extension of municipal piped water supply including fetching water in 20-L plastic jerricans for and extensions to premises | XS | Time saved – fetching water round-trip | Peri-urban | All | Unimproved | 602 | -50.0 | - | -0.395 | Moderate risk | High risk |
| Briand and Lare-Dondarini, (2017) | Burkina Faso | Extension of water infrastructure, through standposts or piped water, and privatised management of the water system | XS with PSM | Time saved – fetching water (average daily collection time) | Rural | All | N/S | 549 | - | -1.34 | -0.209 | Moderate risk | High risk |

|  |  |  |  |  |  |  |  |  |  |  |  | *Risk-of-bias assessment* |  |
| --- | --- | --- | --- | --- | --- | --- | --- | --- | --- | --- | --- | --- | --- |
| *Study* | *Country* | *Intervention detail* | *Study design* | *Outcome* | *Location* | *Participant group* | *Baseline water* | *N* | *mins/trip* | *h/week* | *d* | *Confounding* | *Measurement* |
| Cairncross and Cliff, (1987) | Mozambique | Standpipe provision | Pipeline | Time saved – fetching water (round-trip including queuing time) | Rural | Adult women | Unimproved | 228 | - | -12.4 | -0.707 | Moderate risk | Low risk |
| Cairncross and Cliff (1987) | Mozambique | Standpipe provision | Pipeline | Time spent – household work (including child-care, grinding cereals and food preparation) | Rural | Adult women | Unimproved | 228 | - | 5.72 | 0.327 | Moderate risk | Low risk |
| Cairncross and Cliff (1987) | Mozambique | Standpipe provision | Pipeline | Time spent – working agriculture | Rural | Adult women | Unimproved | 228 | - | 0.70 | 0.040 | Moderate risk | Low risk |
| Cairncross and Cliff (1987) | Mozambique | Standpipe provision | Pipeline | Time spent – leisure (time for eating, personal hygiene and education) | Rural | Adult women | Unimproved | 228 | - | 5.72 | 0.327 | Moderate risk | Low risk |
| Cha et al., (2018) | Ghana | Provision and rehabilitation of boreholes | Cluster-RCT | Time saved – fetching water per day per household | Rural | All | Unimproved | 600 | - | -4.20 | - | Low risk | High risk |
| Dahl-Østergaard et al., (2010) | Yemen | CDD providing dams and dams with irrigation network (pipes or ditches) | DID (XS used in estimation) | Time saved – fetching water per trip | Rural | All – rainy season | Improved | 280 | -30.2 | - | -0.313 | High risk | High risk |
| Dahl-Østergaard et al., (2010) | Yemen | CDD providing private rooftop harvesting tanks | DID (XS used in estimation) | Time saved – fetching water per trip | Rural | All – dry season | Improved | 280 | -39.6 | - | -0.313 | High risk | High risk |
| Dahl-Østergaard et al., (2010) | Yemen | CDD providing water harvesting tanks | DID (XS used in estimation) | Time saved – fetching water per trip | Rural | All – rainy season | Improved | 440 | -1.2 | - | -0.065 | High risk | High risk |
|  |  |  |  |  |  |  |  |  |  |  |  | Confounding | Measurement |
| Dahl-Østergaard et al., (2010) | Yemen | CDD providing piped water | DID (XS used in estimation) | Time saved – fetching water per trip | Rural | All – dry season | Improved | 440 | 29.6 | - | 0.065 | High risk | High risk |
| Dansabo and Gambo, (2019) | Nigeria | Community and Social Development Projects | BA | Time saved | Rural | All adults aged 18+ | Unimproved | 378 |  |  | -0.532 | High risk | High risk |
| David et al. (2004) | St Lucia | Community driven development providing piped water to dwelling/yard and public standpipes | BA | Time saved | Rural | All | Unimproved | 118 | -7.5 | -6.65 | -0.351 | Low risk | High risk |
| Deal and Sabatini, (2020)* | Ghana | Privatisation of local water services: private sector management of improved water from kiosks and boreholes. | CBA (XS used in estimation) | Time saved – fetching water (<30 mins vs >30 mins per trip) | Rural | All | Improved | 1,114 | - | - | 0.096 | High risk | High risk |
| Devoto et al. (2012) | Morocco | Subsidies for piped water connection. | Cluster-RCT (randomised encouragement) | Time spent fetching water over past 3 days in minutes | Rural | All | Improved | 845 |  |  | -0.522 | Low risk | High risk |
| Devoto et al. (2012) | Morocco | Subsidies for piped water connection. | Cluster-RCT (randomised encouragement) | Time spent – leisure over past 7 days (watching TV, exercising, telephone calls, walking, praying, visiting others) | Rural | All | Improved | 845 |  |  | 0.249 | Low risk | High risk |
|  |  |  |  |  |  |  |  |  |  |  |  | Confounding | Measurement |
| Devoto et al. (2012) | Morocco | Subsidies for piped water connection. | Cluster-RCT (randomised encouragement) | Time spent – education (share completing the school year) | Rural | Children | Improved | 363 |  |  | -0.053 | Low risk | High risk |
| Devoto et al. (2012) | Morocco | Subsidies for piped water connection. | Cluster-RCT (randomised encouragement) | Time spent – education (share of completing the school year) | Rural | Girls | Improved | 246 |  |  | 0.051 | Low risk | High risk |
| Devoto et al. (2012) | Morocco | Subsidies for piped water connection. | Cluster-RCT (randomised encouragement) | Time spent – working (female head had income generating activity in past 30 days) | Rural | Females | Improved | 845 |  |  | -0.115 | Low risk | High risk |
| Devoto et al. (2012) | Morocco | Subsidies for piped water connection. | Cluster-RCT (randomised encouragement) | Time spent – working (male head had income generating activity in past 30 days) | Urban | Males | Improved | 845 | - | - | 0.002 | Low risk | High risk |
| Dickinson et al., (2015) | India | CLTS, subsidies and sanitation marketing | Cluster-RCT | Time saved – fetching water (walking time per trip) | Rural | Men | Unimproved | 984 | -3.52 | - | -0.175 | Moderate risk | High risk |
| Dickinson et al. (2015) | India | CLTS, subsidies and sanitation marketing | Cluster-RCT | Time saved – fetching water (walking time per trip) | Rural | Women | Unimproved | 988 | -3.56 | - | -0.200 | Moderate risk | High risk |
| Dickinson et al. (2015) | India | CLTS, subsidies and sanitation marketing | Cluster-RCT | Time saved – fetching water | Rural | Children <5 years | Unimproved | 882 | -2.22 | - | -0.195 | Moderate risk | High risk |
|  |  |  |  |  |  |  |  |  |  |  |  | Confounding | Measurement |
|  |  |  |  | (walking time per trip) |  |  |  |  |  |  |  |  |  |
| Dickinson et al. (2015) | India | CLTS, subsidies and sanitation marketing | Cluster-RCT | Time saved – fetching water (walking time per trip) | Rural | All | Unimproved | 2,854 | -3.10 | - | -0.190 | Moderate risk | High risk |
| Foster et al., (2012) | Kenya | Mobile charging for water utilities | XS regression | Time saved – paying bill (including wait time and return trip) | Urban | All | Improved | 193 | -53.7 | - | -1.595 | High risk | High risk |
| Hasan and Gerber, (2016)* | Bangladesh | Piped improved water provision | XS with PSM | Time saved | Rural | All | Unimproved | 512 | - | -1.09 | -0.374 | High risk | High risk |
| EI Instituto Apoyo, (2000) | Honduras | Community-driven development: water supply improvements | Ex-post pipeline design with group level matching | Time saved | Rural | All | Improved | 1,020 |  | -6.71 | -0.588 | High risk | High risk |
| Instituto Apoyo (2000) | Honduras | Community-driven development: water supply improvements | Ex-post pipeline design with group level matching | Time saved | Rural | Poor people | Improved | 171 |  | -8.46 | -0.926 | High risk | High risk |
| Instituto Apoyo (2000) | Honduras | Community-driven development: water supply improvements | Ex-post pipeline design with group level matching | Time saved | Rural | Extreme poor | Improved | 825 |  | -6.26 | -0.521 | High risk | High risk |
| Jeuland et al., (2015) | India | Community water supply: transportation, treatment and storage of surface water, distributed | DID with PSM | Time saved | Rural | All | Improved | 2,752 | 3.60 | 0.52 | 0.052 | Moderate risk | High risk |
|  |  |  |  |  |  |  |  |  |  |  |  | Confounding | Measurement |
|  |  | by taps at the storage facility |  |  |  |  |  |  |  |  |  |  |  |
| Kumar et al., (2018) | India | SMS notification about water availability | Cluster-RCT | Time saved | Urban | All | Improved | 2,440 | -2.40 |  | -0.025 | High risk | High risk |
| Madajewicz et al., (2007) | Bangladesh | Information about arsenic contamination in public wells used by households | IV design | Time saved | Rural | All | Unimproved | 1,994 | 4.30 |  | 0.420 | Moderate risk | High risk |
| Okyere et al., (2017) | Ghana | Water quality test and information delivered to schools | Cluster-RCT | Time saved – fetching water (round-trip from main drinking water source) | Rural, urban | Adults | Improved | 384 | 0.88 | - | 0.092 | Low risk | High risk |
| Okyere et al. (2017) | Ghana | Water quality test and information delivered to schools | Cluster-RCT | Time saved – fetching water (round-trip from main drinking water source) | Rural, urban | Children (grades 5-8) | Improved | 384 | -3.21 | - | -0.521 | Low risk | High risk |
| Pattanayak et al., (2010) | India | CDD drinking water and sanitation project | Prospective cohort (DID estimation) | Time saved – fetching water (round-trip including waiting time) | Rural | All – rainy season | Unimproved | 1086 | -8 | - | - | Low risk | High risk |
| Pattanayak et al. (2010) | India | CDD drinking water and sanitation project | Prospective cohort (DID estimation) | Time saved – fetching water (round-trip including waiting time) | Rural | All – dry season | Unimproved | 1086 | -1 | - | - | Low risk | High risk |
| Pattanayak et al. (2010) | India | CDD drinking water and sanitation project | Prospective cohort (DID estimation) | Time saved – fetching water (round-trip including waiting time) | Rural | All – rainy and dry seasons | Unimproved | 1086 | -4.5 | - | - | Low risk | High risk |
| Peter, (2010) | Eswatini | Community standpost provision | BA | Time saved – fetching water | Rural | All | Unimproved | 45 | - | - | -2.67 | High risk | High risk |
|  |  |  |  |  |  |  |  |  |  |  |  | Confounding | Measurement |
|  |  |  |  | <30 mins versus 30 mins+ per trip |  |  |  |  |  |  |  |  |  |
| Pories, (2016) | India | Watercredit loans for household connections | BA | Time saved – fetching water (round-trip including waiting time) | Urban | All | Unimproved | 294 | - | -6.3 | - | High risk | High risk |
| Ruben and Zintl, (2011) | Benin | Community water supply and sanitation provision | DID | Time saved | Rural | All | Improved | 3,688 | -14.0 |  | -0.086 | High risk | High risk |
| Ruben and Zintl, (2011) | Benin | Community water supply and sanitation provision | DID with PSM | Time spent – education | Rural | Girls of school age | Improved | 200 | 39.0 |  | 0.332 | High risk | High risk |
| Sakisaka et al., (2015) | Kenya | Installation of tubewells | BA | Time saved – fetching water (median time walking per trip) | - | All | Unimproved | 1,391 | -15.0 | - | -0.181 | Low risk | High risk |
| Schlegel milch et al., (2016) | Kenya | Community water supply and sanitation provision | BA | Time saved – fetching water per trip | Rural | All – rainy season | Unimproved | 250 | -53.5 | - | -0.425 | Moderate risk | High risk |
| Schlegel milch et al. (2016) | Kenya | Community water supply and sanitation provision | BA | Time saved – fetching water per trip | Rural | All – dry season | Unimproved | 250 | -0.5 | - | -0.028 | Moderate risk | High risk |
| Schlegel milch et al., (2016) | Kenya | Community water supply and sanitation provision | BA | Time saved – fetching water per trip | Rural | All – rainy and dry seasons | Unimproved | 250 | -26.5 | - | -0.104 | Moderate risk | High risk |
| Sikder et al., (2020)* | Bangladesh | Chlorination at community hand-pump | XS | Time saved – fetching water (time spent in queue per trip) | Refugee camp | Those living in refugee camp | Unimproved | 19 | 1.50 | - | 2.577 | High risk | High risk |
| Sikder et al. (2020)* | Bangladesh | Chlorination of piped water | XS | Time saved – fetching water (time spent in queue per trip) | Refugee camp | Those living in refugee camp | Unimproved | 20 | 3.9 | - | 1.804 | High risk | High risk |
|  |  |  |  |  |  |  |  |  |  |  |  | Confounding | Measurement |
| Toonen et al., (2014) | Nigeria | Community water supply improvements | XS | Time saved | Rural | Children | Unimproved | 1,105 |  | -2.00 | -0.118 | High risk | High risk |
| WaterAid (2015) | Zambia | New and rehabilitated boreholes in schools and communities | BA | Time saved | Rural | Vulnerable individuals (disability, chronic illness, elderly) | Unimproved | 34 | - | - | -1.036 | High risk | High risk |
| WaterAid (2015) | Zambia | New and rehabilitated boreholes in schools and communities | BA | Time saved | Rural | Vulnerable individuals (disability, chronic illness, elderly) | Unimproved | 34 | - | - | -2.314 | High risk | High risk |
| Winter et al. (2021) | Zambia | Piped water supplies to yards and communal shared areas | DID | Time saved – fetching water (median time to primary source) | Rural | All | Improved | 159 | -15.0 | - | -0.113 | Low risk | Low risk |
| Winter et al. (2021) | Zambia | Piped water supplies to yards and communal shared areas | DID | Time saved – fetching water (all household members) | Rural | All | Improved | 136 | - | -3.80 | -0.577 | Low risk | Low risk |
| Winter et al. (2021) | Zambia | Piped water supplies to yards and communal shared areas | DID | Time saved – fetching water (women household members) | Rural | Women | Improved | 135 | - | -2.90 | -0.579 | Low risk | Low risk |
| Winter et al. (2021) | Zambia | Piped water supplies to yards and communal shared areas | DID | Time saved – fetching water (girl household members) | Rural | Girls | Improved | 41 | - | -1.40 | -0.631 | Low risk | Low risk |
| Winter et al. (2021) | Zambia | Piped water supplies to yards and communal shared areas | DID | Time saved – fetching water (men household members) | Rural | Men | Improved | 18 | - | -0.00 | -0.324 | Low risk | Low risk |
|  |  |  |  |  |  |  |  |  |  |  |  | Confounding | Measurement |
| Winter et al. (2021) | Zambia | Piped water supplies to yards and communal shared areas | DID | Time saved – fetching water (boy household members) | Rural | Boys | Improved | 27 | - | -0.30 | -1.066 | Low risk | Low risk |
| World Bank, (1998) | India (Karnataka) | Water supply improvements, environmental sanitation | BA | Time saved | Rural | All | Improved | 420 | - | -30.6 | - | Low risk | High risk |
| World Bank (1998) | India (Maharashtra) | Water supply improvements, environmental sanitation | BA | Time saved | Rural | All | Improved | 589 | - | -16.1 | - | Low risk | High risk |
| Ziegelhofer, (2012) | Guinea | Community water supply providing and rehabilitating water wells, and solar-based systems | RDD | Time saved – fetching water per day | Rural | All – rainy season | Unimproved | 1,139 | - | -2.17 | -0.369 | Low risk | High risk |
| Ziegelhofer (2012) | Guinea | Community water supply providing and rehabilitating water wells, and solar-based systems | RDD | Time saved – fetching water per day | Rural | All – rainy and dry seasons | Unimproved | 1,139 | - | -3.52 | -0.541 | Low risk | High risk |
| Ziegelhofer (2012) | Guinea | Community water supply providing and rehabilitating water wells, and solar-based systems | RDD | Time saved – fetching water per trip | Rural | All – dry season | Unimproved | 1,139 | -10.9 | - | -0.666 | Low risk | High risk |
| Ziegelhofer (2012) | Guinea | Community water supply providing and rehabilitating water wells, and solar-based systems | RDD | Time saved – fetching water per trip | Rural | All – rainy season | Unimproved | 1,139 | -3.58 | - | -0.260 | Low risk | High risk |
|  |  |  |  |  |  |  |  |  |  |  |  | Confounding | Measurement |
| Ziegelhofer (2012) | Guinea | Community water supply providing and rehabilitating water wells, and solar-based systems | RDD | Time saved – fetching water per trip | Rural | All – rainy and dry seasons | Unimproved | 1,139 | -7.23 | - | -0.481 | Low risk | High risk |

Notes: \* water treatment interventions: treated water sold at kiosks (Deal, 2020), piped chlorinated water (Hasan, 2016), chlorination at water source (Sikder, 2020), information about water quality (Okyere et al. (2017) and information about arsenic (Madajewicz et al. (2007). BA before after design; CDD community-driven development; DID difference in differences, IV instrumental variables; PSM propensity score matching; RDD regression discontinuity design; XS cross section design. Counterfactual baseline scenarios defined by JMP (WHO/UNICEF, 2023).

**Table A2.** Sanitation interventions.

| Study | Country | Intervention detail | Study design | Outcome measure | Participant group | N | mins/trip | h/week | d | Risk-of-bias assessment |  |
| --- | --- | --- | --- | --- | --- | --- | --- | --- | --- | --- | --- |
|  |  |  |  |  |  |  |  |  |  | Confounding | Measurement |
| Biran et al., (2018) | Malawi | Disability-inclusive CLTS | Cluster-RCT | Time saved walking to toilet in minutes | People with disabilities aged >2 | 171 | -0.2 | - | -0.036 | Low risk | Low risk |
| Cha et al., (2020) | Ethiopia | CLTS with hand-washing stations | Cluster-RCT | Time saved walking to place of defaecation hours per household | 5-14s | 3,972 | - | -5.6 | -0.382 | Low risk | High risk |
| Cha et al. (2020) | Ethiopia | CLTS with hand-washing stations | Cluster-RCT | Time saved walking to place of defaecation hours per household | Over-15s | 4,776 | - | -4.0 | -0.175 | Low risk | High risk |
| Cha et al. (2020) | Ethiopia | CLTS with hand-washing stations | Cluster-RCT | Time saved walking to place of defaecation hours per household | Over-5s | 8,580 | - | -4.7 | -0.209 | Low risk | High risk |
| Cha et al. (2020) | Ethiopia | CLTS with hand-washing stations | Cluster-RCT | Time saved from being sick or caring for sick hours per household | Under-5s | 1,469 | - | -2.4 | - | Low risk | High risk |
| Cha et al. (2020) | Ethiopia | CLTS with hand-washing stations | Cluster-RCT | Time saved from being sick or caring for sick hours per household | 5-14s | 3,972 | - | -0.7 | - | Low risk | High risk |
| Cha et al. (2020) | Ethiopia | CLTS with hand-washing stations | Cluster-RCT | Time saved from being sick or caring for sick hours per household | Over-15s | 4,776 | - | -0.5 | - | Low risk | High risk |
| Cha et al. (2020) | Ethiopia | CLTS with hand-washing stations | Cluster-RCT | Time saved from being sick or caring for sick hours per household | All | 9,881 | - | -0.8 | - | Low risk | High risk |
| Cha et al. (2020) | Ethiopia | CLTS with hand-washing stations | Cluster-RCT | Time saved hours per household | 5-14s | 3,972 | - | -6.2 | - | Low risk | High risk |
| Cha et al. (2020) | Ethiopia | CLTS with hand-washing stations | Cluster-RCT | Time saved hours per household | Over-15s | 4,776 | - | -4.5 | - | Low risk | High risk |
| Cha et al. (2020) | Ethiopia | CLTS with hand-washing stations | Cluster-RCT | Time saved hours per household | All | 9,881 | - | -4.9 | - | Low risk | High risk |
| Dickinson et al. (2015) | India | CLTS, subsidies and | Cluster-RCT | Time saved walking to place of defaecation mins | Men | 984 | -3.5 | - | -0.175 | Moderate risk | High risk |
|  |  |  |  |  |  |  |  |  |  | Confounding | Measurement |
|  |  | sanitation marketing |  |  |  |  |  |  |  |  |  |
| Dickinson et al. (2015) | India | CLTS, subsidies and sanitation marketing | Cluster-RCT | Time saved walking to place of defaecation mins | Women | 988 | -3.6 | - | -0.200 | Moderate risk | High risk |
| Dickinson et al. (2015) | India | CLTS, subsidies and sanitation marketing | Cluster-RCT | Time saved walking to place of defaecation mins | Children <5 years | 882 | -2.2 | - | -0.195 | Moderate risk | High risk |
| Dickinson et al. (2015) | India | CLTS, subsidies and sanitation marketing | Cluster-RCT | Time saved walking to place of defaecation mins | All | 2,854 | -3.1 | - | -0.190 | Moderate risk | High risk |
| Hutton et al. (2020) | India | CLTS, subsidies and sanitation marketing | Before versus after | Time saved per person from using toilet in hours per household | All | 10,051 | - | -3.8 | - | High risk | High risk |
| Pattanayak et al., (2010) | India | CDD - rural drinking water and sanitation project | Prospective cohort (DID estimation) | Times saved walking to place of defaecation in minutes | All | 1,086 | -4.7 | - | -0.159 | Low risk | High risk |
| WaterAid (2015) | Zambia | Disability-friendly CLTS | Before versus after | Time saved to walk to latrine | Vulnerable individuals (disability, chronic illness, elderly) | 34 | - | - | -1.036 | Moderate risk | High risk |

Notes: CDD community-driven development; CLTS community led total sanitation; DID difference in differences. All studies were conducted of latrine provision or promotion in rural areas and used counterfactual scenarios where most households used sanitation that was 'unimproved' as defined by JMP (WHO/UNICEF, 2023).

**Table A3.**
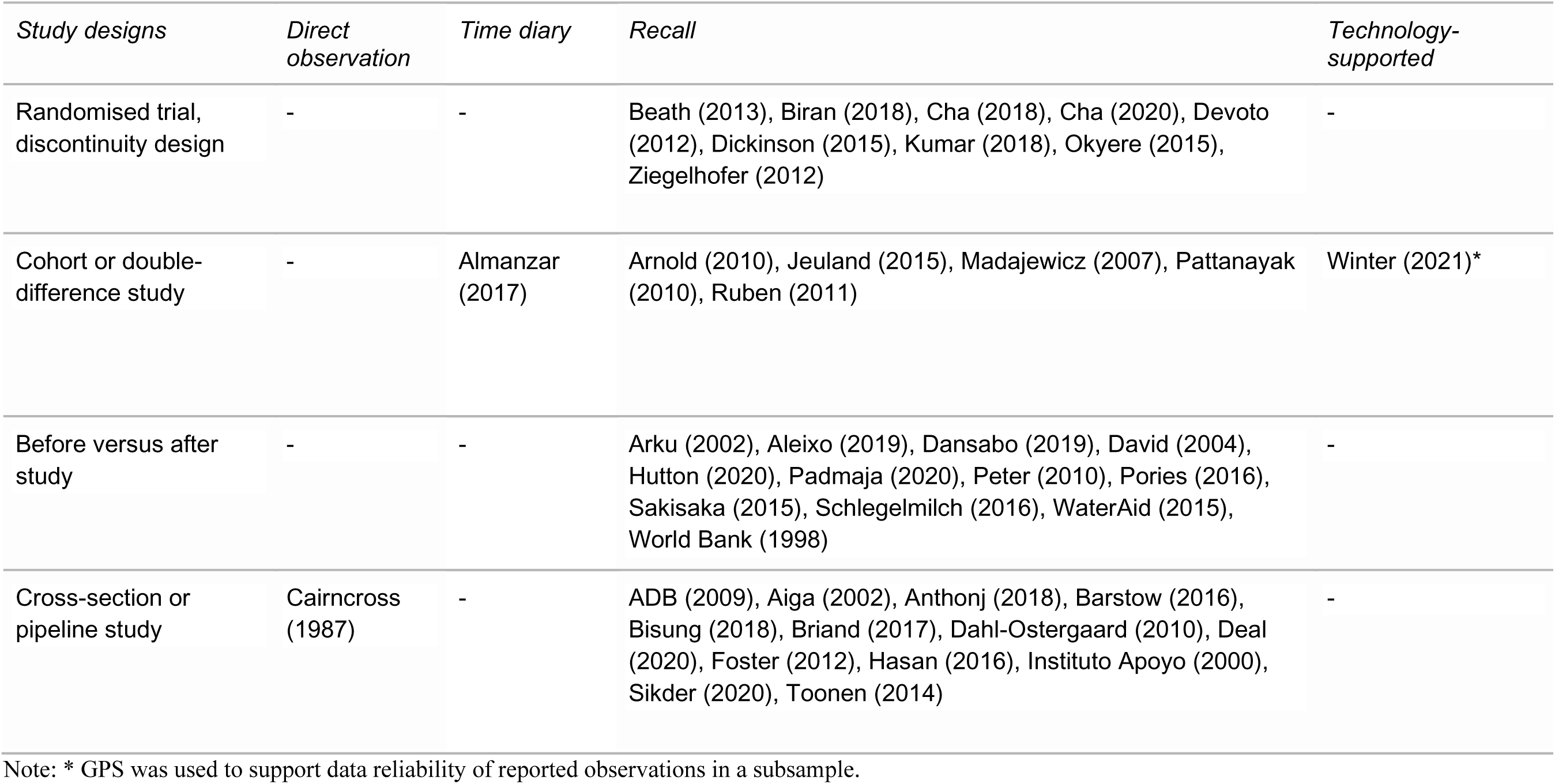
Study designs used to evaluate the impact of WASH interventions on travel time and time use.

### Supplementary Annex 3: Additional analysis

**Table A4.** Change in time associated with WASH improvements by global region.

| *Intervention* | *Outcome* | *Mean* | *Median* | *SD* | *Minimum* | *Maximum* | *Number of estimates* |
| --- | --- | --- | --- | --- | --- | --- | --- |
| Water supply (LAC) | Minutes per trip | -6.1 | -5.0 | 6.1 | -14.0 | -0.4 | 4 |
|  | Hours per week | -5.3 | -6.3 | 2.3 | -8.5 | -2.2 | 7 |
| Water supply (SEA) | Minutes per trip | -4.0 | -3.5 | 3.0 | -8.0 | -1.0 | 4 |
|  | Hours per week | -11.0 | -2.9 | 14.0 | -36.1 | -0.2 | 15 |
| Water supply (SSA) | Minutes per trip | -21.2 | -3.2 | 25.5 | -66.4 | 0.9 | 13 |
|  | Hours per week | -4.4 | -3.5 | 3.8 | -12.4 | -1.3 | 7 |
| Sanitation (LAC) | Minutes per trip | - | - | - | - | - | 0 |
|  | Hours per week | - | - | - | - | - | 0 |
| Sanitation (SWA) | Minutes per trip | -3.4 | -3.5 | 0.9 | -4.7 | -2.2 | 5 |
|  | Hours per week | -3.8 | -3.8 | - | -3.8 | -3.8 | 1 |
| Sanitation (SSA) | Minutes per trip | -0.2 | -0.2 | - | -0.2 | -0.2 | 1 |
|  | Hours per week | -3.4 | -4.3 | 2.2 | -6.2 | -0.5 | 10 |

Notes: values of mean < 0 indicate reduction in travel and access time (a time saving) following intervention; LAC Latin American and Caribbean; SEA South and East Asia; SWA South and Western Asia; SSA sub-Saharan Africa; SD standard deviation.

#### Publication bias assessment

Publication bias tests suggested some evidence for small study effects for travel and access time outcomes. The funnel graph presenting the distribution of effects and standard errors, together with the regression line, appeared clearly asymmetrical for travel time in the region of underpowered studies, consistent with the possibility of publication bias (Figure A1). Egger et al. (1998) test coefficients associated with small study effects were of the expected sign and statistically significant (intercept=-1.12, 95%CI=-1.97, -0.29; 44 observations). The asymmetry was less clear in the funnel graph for time use estimates (Figure A2) and the regression coefficient was neither of the expected sign (given that alternate time use outcomes are expected to be positive rather than negative) nor statistically significant (intercept=-0.62, 95%CI=-1.38, -0.14; 34 observations). These findings were supported in the meta-regression analysis, which found significantly smaller magnitude effects in studies published in journals, suggesting direct evidence for publication bias for travel time estimates.

**Figure A1.**
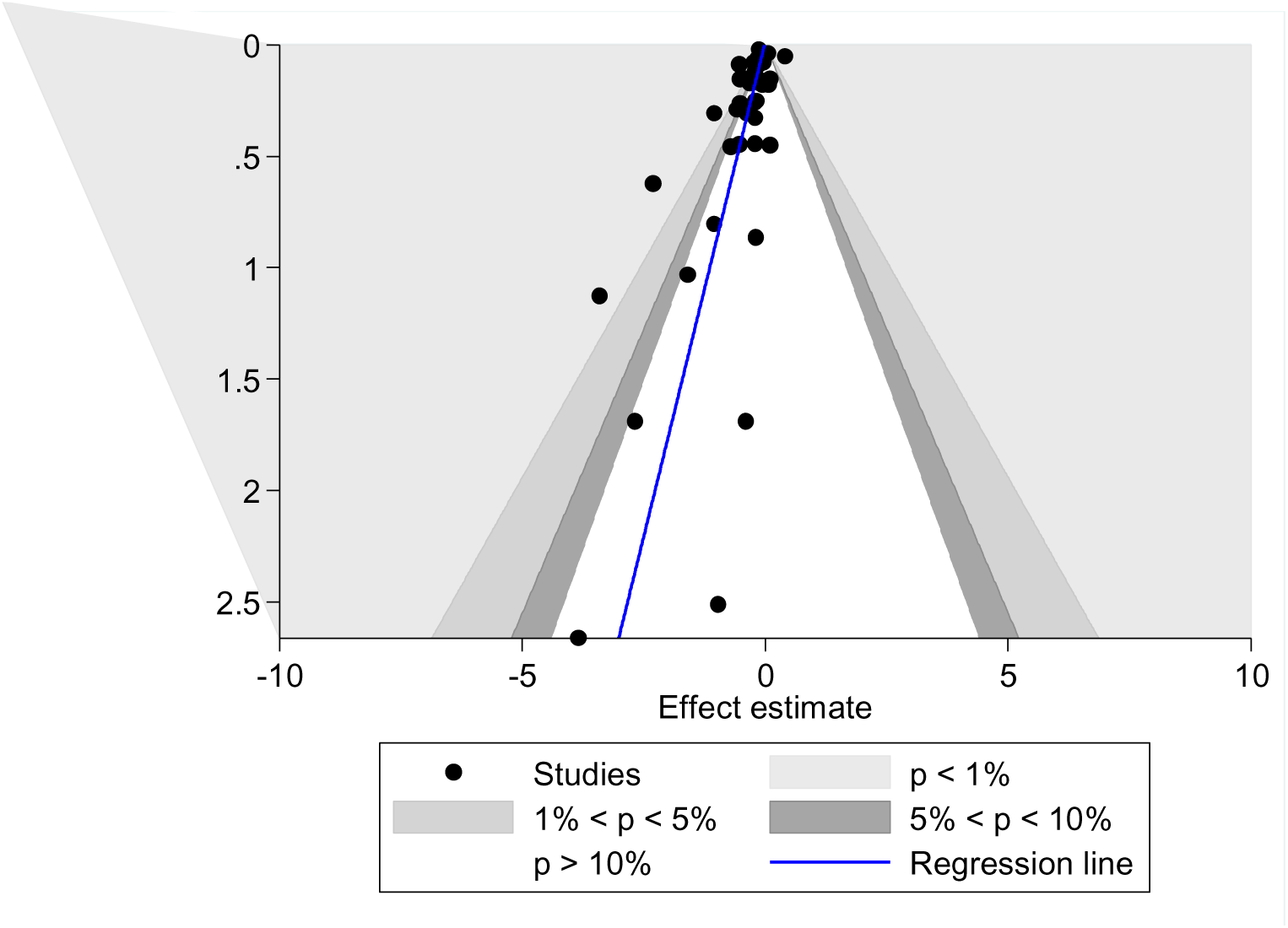
Funnel graph with regression lines for travel and access time outcomes

**Figure A2.**
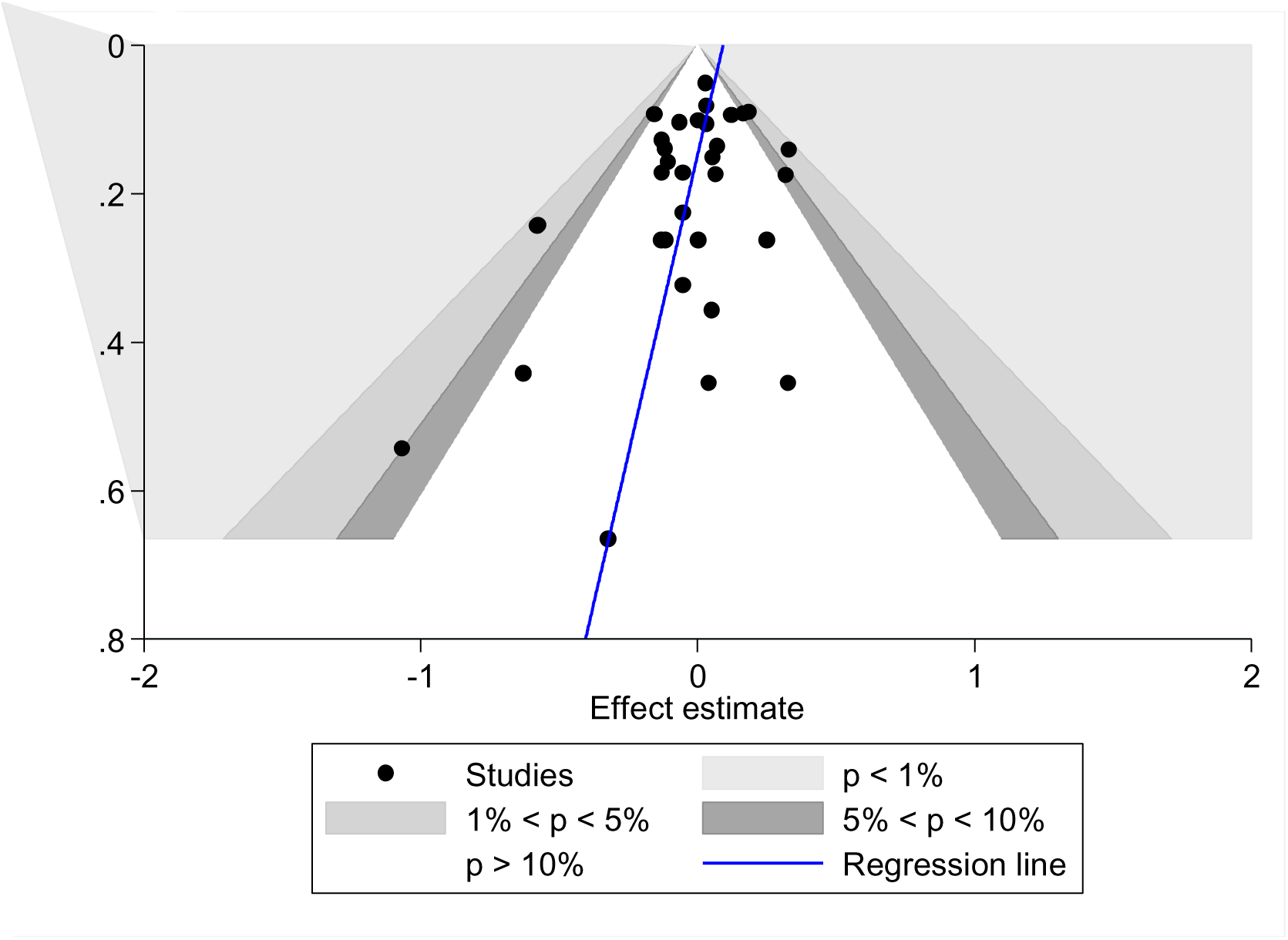
Funnel graph with regression lines for alternative time use outcomes

### Meta-regression analysis

We attempted to explain the heterogeneity in findings using meta-regression. We split the analysis between travel time and time reallocation outcomes, because they represented different stages of the causal pathway in the theory of change (Figure 1). The analysis for travel and access time suggested that water treatment and quality interventions as a group were associated with significantly increased time burdens over the measured counterfactuals (Table A6). The interventions with the largest effects on reducing travel and access time were those providing mobile billing (m-WASH) interventions. Publication year was included as a measure of general progress in providing access to WASH services; effects on both travel and access time and time use reallocation were absolutely smaller in more recent studies, which tend to be conducted under relatively improved counterfactual WASH scenarios. Interventions for vulnerable populations (disabled, chronically ill and elderly) tended to have significantly larger effects on reducing travel and access time. Effects measured in hours per week also tended to be of significantly larger magnitude than those measured in minutes per trip, which is indicative of the greater time savings over multiple trips per day and household members. For time use reallocation, three coefficients were statistically significantly different from zero, indicating larger effects of water supply interventions and effects measured among children, and smaller effects for more recent publications (Table A6).

**Table A6:** Meta-regression analysis results: travel time and time reallocation.

|  | Travel time |  |  |  | Time use |  |  |  |
| --- | --- | --- | --- | --- | --- | --- | --- | --- |
|  | Coeff | P>t | 95%CI L | 95%CI U | Coeff | P>t | 95%CI L | 95%CI U |
| Water treatment/quality | 0.359** | 0.001 | 0.164 | 0.555 |  |  |  |  |
| Water supply |  |  |  |  | 0.460** | 0.02 | 0.090 | 0.829 |
| CDD+ |  |  |  |  |  |  |  |  |
| CLTS | -0.230 | 0.202 | -0.589 | 0.129 |  |  |  |  |
| Community water supply | -0.180** | 0.016 | -0.325 | -0.036 |  |  |  |  |
| m-WASH | -0.408** | 0.020 | -0.749 | -0.068 |  |  |  |  |
| Piped water supply | -0.173 | 0.281 | -0.495 | 0.148 |  |  |  |  |
| Year of publication | 0.015* | 0.080 | -0.002 | 0.032 | -0.012* | 0.07 | -0.025 | 0.001 |
| General population+ |  |  |  |  |  |  |  |  |
| Vulnerable | -0.720** | 0.011 | -1.265 | -0.175 |  |  |  |  |
| Child |  |  |  |  | 0.127** | 0.03 | 0.013 | 0.242 |
| Hours per week |  |  |  |  |  |  |  |  |
| measure+ |  |  |  |  |  |  |  |  |
| Minutes per trip measure | 0.219*** | 0.004 | 0.074 | 0.364 |  |  |  |  |
| High risk of bias | -0.564* | 0.066 | -1.167 | 0.038 |  |  |  |  |
| Low/medium risk of |  |  |  |  |  |  |  |  |
| bias+ |  |  |  |  |  |  |  |  |
| Small study effect |  |  |  |  |  |  |  |  |
| (standard error of d) | -1.288*** | 0.000 | -1.869 | -0.708 | -0.422 | 0.29 | -1.215 | 0.371 |
| Published in journal | 0.213* | 0.052 | -0.002 | 0.428 |  |  |  |  |
| Constant | 0.100 | 0.753 | -0.540 | 0.739 | -0.094 | 0.77 | -0.740 | 0.552 |
| Number of observations | 44 |  |  |  | 34 |  |  |  |
| Tau-squared | 0.007 |  |  |  | 0.000 |  |  |  |
| I-squared | 7% |  |  |  | 0% |  |  |  |
| Adjusted R-squared | 84% |  |  |  | 100% |  |  |  |
| Model F | 7.35 |  |  |  | 8.46 |  |  |  |
| Prob > F | 0.000 |  |  |  | 0.000 |  |  |  |

Notes: + reference category; \*\*\*, \*\*, \* indicate coefficient is statistically significant at p<0.01, p<0.05,
p<0.1.

## Footnotes

1 The data for travel time in hours per week were very skewed; the mean travel and access time savings were 4.5 hours in sub-Saharan Africa, 5.5 hours in Latin America and 11 hours in South and East Asia.

2 The five biggest effect sizes were calculated from dichotomous measures. Peter et al. (2010) measured time savings (<30 mins versus 30+ mins) from installation of a community standpost in rural Swaziland (now Eswatini). WaterAid (2015) measured time savings (<30 mins versus 30+ mins travel time) among vulnerable individuals. Arku (2002) measured savings as under one-hour versus 1 or more hours in Ghana. Foster et al. (2012) measured time savings from mobile water tariff payments versus payment at the bank (including wait time and return trip) in urban Kenya. Aleixo et al. (2019) measured time savings from household piped water connections as those with zero time spent versus those spending time.

